# A Streamlined Point-of-Care CRISPR Test for Tuberculosis Detection Directly from Sputum

**DOI:** 10.1101/2025.02.19.25322517

**Authors:** Owen R. S. Dunkley, Alexandra G. Bell, Nisha H. Modi, Yujia Huang, Soleil Tseng, Robert Reiss, Naranjargal Daivaa, J. Lucian Davis, Deninson Alejandro Vargas, Padmapriya Banada, Yingda L. Xie, Cameron Myhrvold

**Affiliations:** Department of Molecular Biology, Princeton University, Princeton New Jersey, 08544, USA; Public Health Research Institute, Department of Medicine, Rutgers New Jersey Medical School, Newark, New Jersey, 07103, USA; Department of Epidemiology of Microbial Diseases, Yale School of Public Health, New Haven, Connecticut, United States of America; Pulmonary, Critical Care, and Sleep Medicine Section, Yale School of Medicine, New Haven, Connecticut, United States of America; Centro Internacional de Entrenamiento e Investigaciones Médicas (CIDEIM), Cali, Colombia; Universidad Icesi, Cali, Colombia; Department of Chemical and Biological Engineering, Princeton University, Princeton, New Jersey, 08544, USA; Omenn-Darling Bioengineering Institute, Princeton University, Princeton, New Jersey, 08544, USA; Department of Chemistry, Princeton University, Princeton, New Jersey, 08544, USA

**Author notes:** Department of Bioengineering, Stanford University, Stanford, California 94305, USA. These authors contributed equally (Owen R. S. Dunkley, Alexandra G. Bell). These authors contributed equally (Padmapriya Banada, Yingda L. Xie, Cameron Myhrvold).

## Abstract

*Mycobacterium tuberculosis* (*Mtb*) is a major threat to global health and is responsible for over one million deaths each year. To stem the tide of cases and maximize opportunities for early interventions, there is an urgent need for affordable and simple means of tuberculosis diagnosis in under-resourced areas. We sought to develop a CRISPR-based isothermal assay coupled with a compatible, straightforward sample processing technique for point-of-care use. Here, we combine Recombinase Polymerase Amplification (RPA) with Cas13a and Cas12a, to create two parallelised one-pot assays that detect two conserved elements of *Mtb* (*IS6110* and *IS1081*) and an internal control targeting human DNA. These assays were shown to be compatible with lateral flow and can be readily lyophilized. Our finalized assay exhibited sensitivity over a wide range of bacterial loads (10^5^ to 10^2^ CFU/mL) in sputum. The limit of detection (LoD) of the assay was determined to be 69.0 (51.0 – 86.9) CFU/mL for *Mtb* strain H37Rv spiked in sputum and 80.5 (59.4 – 101.6) CFU/mL for *M. bovis* BCG. Our assay showed no cross reactivity against a wide range of bacterial/fungal isolates. Clinical tests on 13 blinded sputum samples revealed 100% (6/6) sensitivity and 100% (7/7) specificity compared to culture. Our assay exhibited comparable sensitivity in clinical samples to the microbiological gold standard, TB culture, and to the nucleic acid state-of-the-art, GeneXpert MTB/RIF Ultra. This technology streamlines TB diagnosis from sample extraction to assay readout in a rapid and robust format, making it the first test to combine amplification and detection while being compatible with both lateral flow and lyophilization.

## Introduction

Tuberculosis (TB), caused by the pathogen *Mycobacterium tuberculosis* (*Mtb*), is a major threat to global health and is difficult to diagnose^1^. Current TB diagnostic methods are limited by trade-offs among sensitivity, turnaround time, and accessibility. Culture-based methods are very sensitive, but the turnaround time is slow (weeks)^2^. Conversely, sputum smear microscopy is rapid (∼ 1 hour turnaround) but has limited sensitivity and specificity, in addition to relying on user expertise^3–5^. PCR-based tests, such as GeneXpert MTB/RIF, are sensitive but they require specialized equipment with an integrated thermocycler and proprietary single-use cartridges^6–8^. Thus, there remains a critical unmet need for sensitive point-of-care TB diagnostics in low-resourced areas.

CRISPR-based diagnostics (CRISPR-Dx) have the potential to bridge the accessibility and accuracy gap by leveraging the high specificity and collateral cleavage activities of CRISPR-associated proteins such as Cas12 and Cas13 and pairing them with isothermal amplification techniques^9–16^. Upon target recognition, Cas12 and Cas13 cleave their targets in *cis* and cleave nearby single-stranded DNA or RNA, respectively, in *trans*^*14*,*15* ,17^. Thus, by including a quenched fluorescent reporter with a cleavable nucleic acid linker, target recognition can be converted to a fluorescent signal^16^. Cas12 and Cas13 require a high-degree of CRISPR RNA (crRNA)-target complementarity to activate their nuclease domains, making reporter cleavage highly specific to the presence of a target sequence^14,17,18^. Two representative subtypes, Cas12a and Cas13a, both exhibit high turnover catalytic activity, and pairing them with an excess of cleavable nucleic acid reporter provides improved signal amplification^9,19,14,16,20,21^. When combined with isothermal amplification, these properties enable robust, highly specific detection with clinically relevant sensitivity—without the need for thermocycling^9,10,14,22^.

Several groups have employed CRISPR-Dx paired with PCR or isothermal amplification to address accessibility issues in TB diagnosis^23–28^. However, these methods require user manipulations and specialized instrumentation between the preamplification of the sample and detection, making these assays hard to deploy and exposing the workflow to potential sources of contamination. Peng and colleagues developed a Cas12-based format that combines amplification and detection but requires instrument-based nucleic acid extraction and readout^22,29^. Therefore, there is an unmet need for CRISPR-Dx methods designed to work in constrained environments with simplified sample preparation techniques for the detection of tuberculosis-causing mycobacteria without specialized instrumentation.

In this study, we have developed SHINE-TB from our general-purpose diagnostic platform, SHINE (Streamlined Highlighting of Infections to Navigate Epidemics)^12,13^. SHINE-TB consists of two parallelised single-pot reactions that combine isothermal amplification by Recombinase Polymerase Amplification (RPA)^30^, *in vitro* transcription, and detection, including a Cas13a assay for detecting two conserved elements in the *Mtb* genome and a Cas12a assay for detecting human DNA as an internal control. Combined with an efficient sample processing method and an optimized CRISPR Cas13a/Cas12a assay, we present a simple, point-of-care oriented, rapid and highly sensitive TB detection system that requires no expensive instrumentation^31^.

## Results

### One-Pot Cas12a Assay Accurately Detects *Mtb* at Genome-Level Sensitivity

We first explored a streamlined Cas12a-based diagnostic workflow for *Mtb*, where isothermal amplification of the DNA target using RPA is paired with direct detection by *Lachnospiraceae bacterium* Cas12a (*Lb*Cas12a enzyme, referred to herein as Cas12a). A Cas12a workflow permits using fewer reaction components than a Cas13a workflow because the enzyme can directly detect the RPA product, eliminating the need for *in vitro* transcription (Fig. 1a). We began by designing and testing a set of six RPA primer and Cas12a crRNA sets, targeting the multicopy insertion sequences *IS6110* and *IS1081*, found uniquely within the member species of the *Mtb* complex (MTBC) (for details, see Methods).

**Figure 1.**
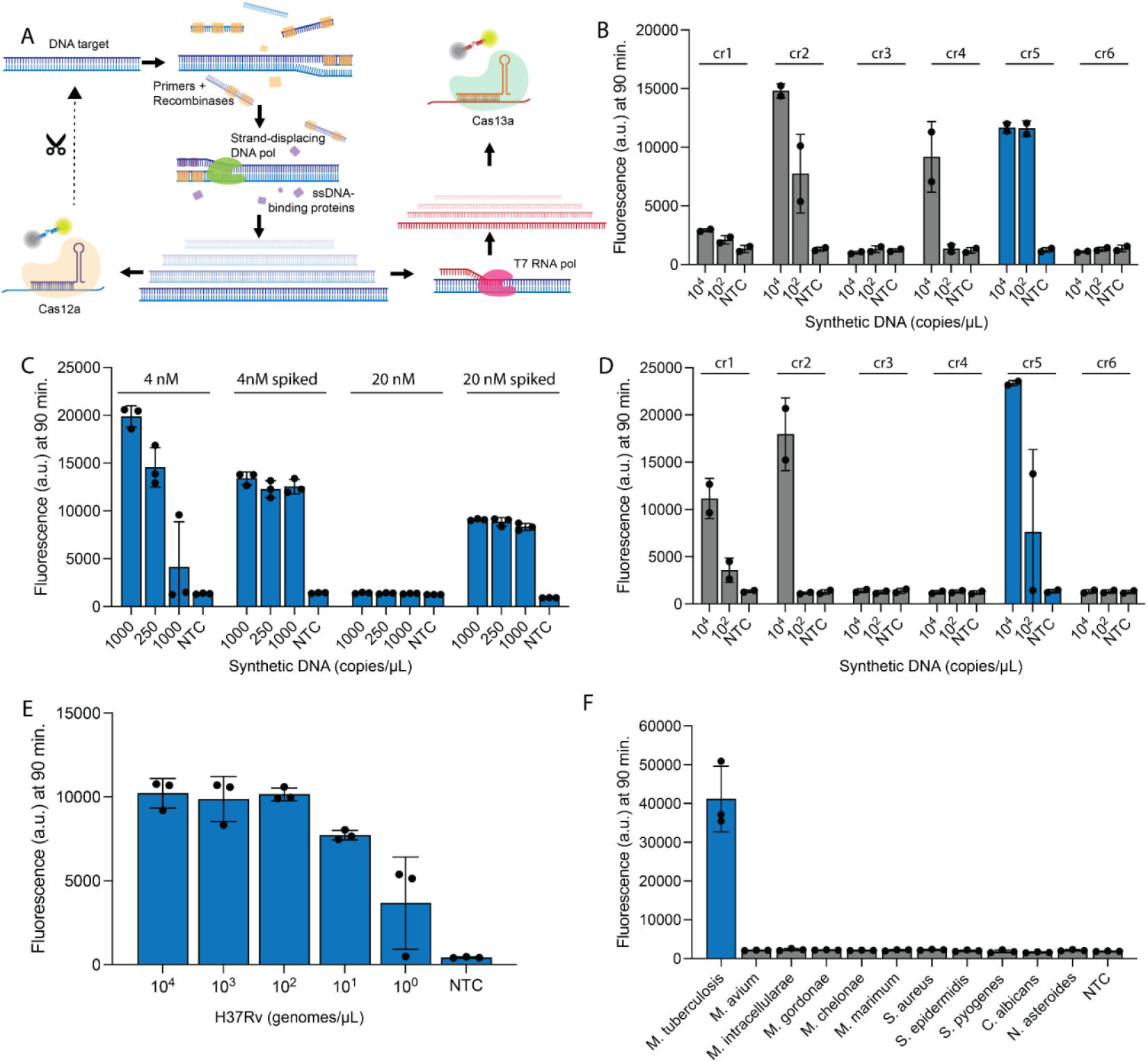
CRISPR-Cas12a assay optimization for detecting *Mtb*. A. Schematic of Cas-12a/Cas13a based CRISPR reaction, where DNA is amplified using target specific primers, recombinases, strand-displacing polymerase, and ssDNA binding proteins and directly detected by Cas12a or transcribed to RNA with T7 reverse transcription for detection with Cas13a. **B**. Cas12a detecting synthetic DNA for six different crRNA-primer designs. Amplification was allowed to proceed for 30 minutes before the addition of crRNA and Cas12a. **C**. Cas12a detecting synthetic DNA with varying concentrations of Cas12a. Molar concentration alone indicates all components present from the start of the reaction. Molar concentration spiked indicates amplification was allowed to proceed for 30 minutes before the addition of crRNA and Cas12a. **D**. Cas12a detecting synthetic DNA in a one-pot format, as all components are present from the start of the reaction. **E**. Performance of Cas12a assay for decreasing concentrations of H37Rv genome. **F**. Specificity panel for Cas12a assay against various NTMs and other bacteria/fungi at 1 ng/µL. In B and D, error bars: SD based on n=3 replicates. In C, E, and F, error bars: SD based on n=2 replicates.

To compare the sensitivity of each design for amplification and detection as isolated events, we carried out RPA for 30 minutes before spiking Cas12a into the reactions (Fig. 1b and Supplementary Fig. 1a). Primer-crRNA designs cr2 and cr5 outperformed our other assays, detecting 100 copies of the synthetic insertion sequence per µL of the input synthetic DNA (Fig. 1b). As *cis* cleavage of the target amplicon in a one-pot reaction interferes with further amplification^32,33^, we aimed to maximize the potential for RPA to overcome the rate of Cas12a *cis* cleavage. We varied the concentration of Cas12a in one-pot and spiked-enzyme conditions targeting synthetic copies of the insertion sequence, finding that lower concentrations of Cas12a improved the sensitivity of the one-pot assay (Fig. 1c and Supplementary Fig. 1b). Design cr5 appeared to be the most effective combination of primers and a crRNA in these improved conditions for one-pot detection targeting the synthetic insertion sequence (Fig. 1d and Supplementary Fig. 2a). Even with conditions modified to improve assay sensitivity, the one-pot format still performed less sensitively than did the initial spike-in experiments, as seen for designs cr2, cr4, and cr5, possibly due to incomplete amplification (Fig. 1b and Fig. 1d).

The analytical sensitivity of the Cas12a assay (design cr5) was determined using H37Rv genomic DNA, and specificity was evaluated against 10 nontuberculous mycobacteria (NTMs) and other pathogens. The H37Rv genome contains 17 copies of *IS6110*, and despite the added complexity of the genomic material as compared to synthetic copies of the insertion sequence, our one-pot Cas12a cr5 assay could detect H37Rv DNA at 10 genomes/µL (∼170 copies/µL of *IS6110*) in 100% (3/3) of the replicates and could detect down to 1 genome/µL in 2 out of 3 (66%) technical replicates (Fig. 1e and Supplementary Fig. 2b). Design cr5 showed no cross-reactivity with NTMs and other pathogens at 1 ng/µL per test (Fig. 1f and Supplementary Fig. 2c).

### Cas13a Assay for *Mtb* Detection Overcomes Cas12a Limitations

We reasoned that the decreased sensitivity of the Cas12a-based assays in a one-pot format seen in Fig. 1d compared to Fig. 1b was due to Cas12a cleaving the target DNA in *cis* before sufficient amplification could occur, thereby reducing exponential target amplification and *trans* cleavage of the reporter (Fig. 1a). As Cas13a requires an RNA target and cleaves RNA in *trans*, we hypothesized that Cas13a would provide higher sensitivity in a single pot reaction as it would not disrupt amplification in the same manner as Cas12a despite requiring the added complexity of *in vitro* transcription in the one-pot reaction.

To test this hypothesis, we used Activity-informed Design with All-inclusive Patrolling of Targets (ADAPT) to design six different crRNA sets for *Leptotrichia wadei* (*Lwa*) Cas13a (herein referred to as Cas13a) for two high copy number insertion sequences, three targeting *IS6110* and three targeting *IS1081*^*34*^. We experimentally tested each design with synthetic targets in a one-pot format, identifying IS6110c and IS1081a as promising, as both detected 2 of 3 replicates at 10 copies/µL and all higher concentrations (Fig. 2a, Supplementary Fig. 3a). Head-to-head comparison using serial dilutions of synthetic targets in Cas12a- and Cas13a-based one-pot *IS6110* assays showed that Cas13a IS6110c was more sensitive at detection limits of 1 copy/µL of *IS6110*, whereas Cas12a cr5 began to lose signal at 384 copies/µL (Fig. 2b and Supplementary Fig. 3b). This demonstrates that Cas13a has higher sensitivity compared to Cas12a when amplification is combined with detection in a single pot. In subsequent experiments, we used Cas13a for *Mtb* detection.

**Figure 2.**
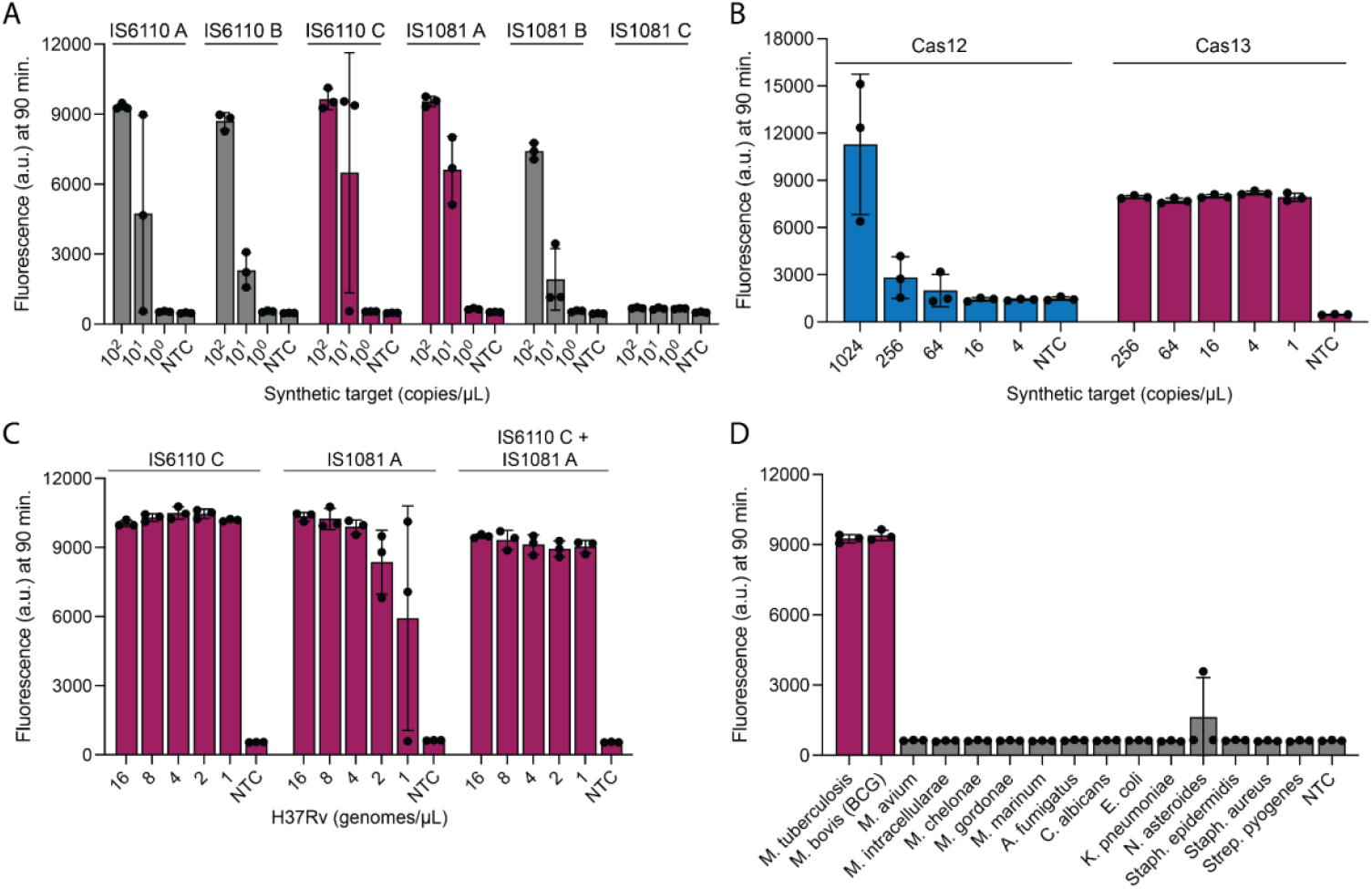
CRISPR-Cas13a based detection of *Mtb*. A. Cas13a detecting synthetic targets for six different crRNA-primer designs against two different targets, IS6110 and IS1081. **B**. Comparison of Cas12a to Cas13a detection of IS6110 synthetic target. Cas13a is a single-crRNA assay for direct comparison. **C**. Cas13a detecting H37Rv in single-target detection (IS6110 only) and dual-target detection (IS6110 and IS1081) formats. **D**. Cross reactivity panel for Cas13a dual detection assay against various NTMs and other pathogens at 0.1 ng/µL. Error bars: SD based on n=3 replicates.

We further improved the sensitivity of the assay by targeting both *IS6110* and *IS1081* in a dual detection assay (Fig. 2c). Combining IS1081a and IS6110c into one reaction did not appear to hamper either assay’s sensitivity (Fig. 2c and Supplementary Fig. 3c). There was no significant cross-reactivity of the combined *IS6110* and *IS1081* dual detection assay against 13 NTMs and other pathogens at 0.1 ng/µL (Fig. 2d and Supplementary Fig. 3d). One replicate for *N. asteroides* had fluorescence above background but upon retesting this pathogen with 2.5 ng/µL of input there was no signal, suggesting that the assay is highly specific (Supplementary Fig. 3e).

### Adapting Dual Detection CRISPR Assay (IS6110+IS1081) for Clinical Settings

We incorporated an internal control targeting human DNA into our diagnostic system to 1) to avoid reporting false negative results due to potential interference from sample inhibitors or poor nucleic acid extraction etc., and 2) as sample adequacy control, where it ensures the target nucleic acid was from a human derived sample. After evaluating several targets, we included a Cas12a-based internal control that targets the Long Terminal Repeat (LTR) of the endogenous retrovirus ERVK, as each human chromosome carries between 40 and 400 copies of the element^35^ (Supplementary Fig. 4). Due to the target’s abundance in human DNA, we were able to rely on detection without reducing the concentration of Cas12a for our optimized one-pot assay (Supplementary Fig. 4), even for challenging sample types.

To enable SHINE testing of sputum, the standard sample type for pulmonary TB diagnosis, we used a novel sample processing method developed in parallel by Modi *et al*^*31*^. This method processes sputum samples for detection by our CRISPR assays and other nucleic acid amplification methods (see methods) while circumventing the need for specialized equipment^31^. We tested the compatibility of our assay with sputum by spiking BCG genomic DNA at known concentrations into pooled and diluted sputum samples collected from TB-negative patients at University Hospital in Newark, NJ. As low as 10^2^ copies of the BCG genome per µL of sputum (the lowest tested concentration in this experiment) were consistently detected by both the Cas12a internal control and Cas13a dual detection assay, indicating compatibility with this clinically relevant sample type (Fig. 3a and Supplementary Fig. 5a).

**Figure 3.**
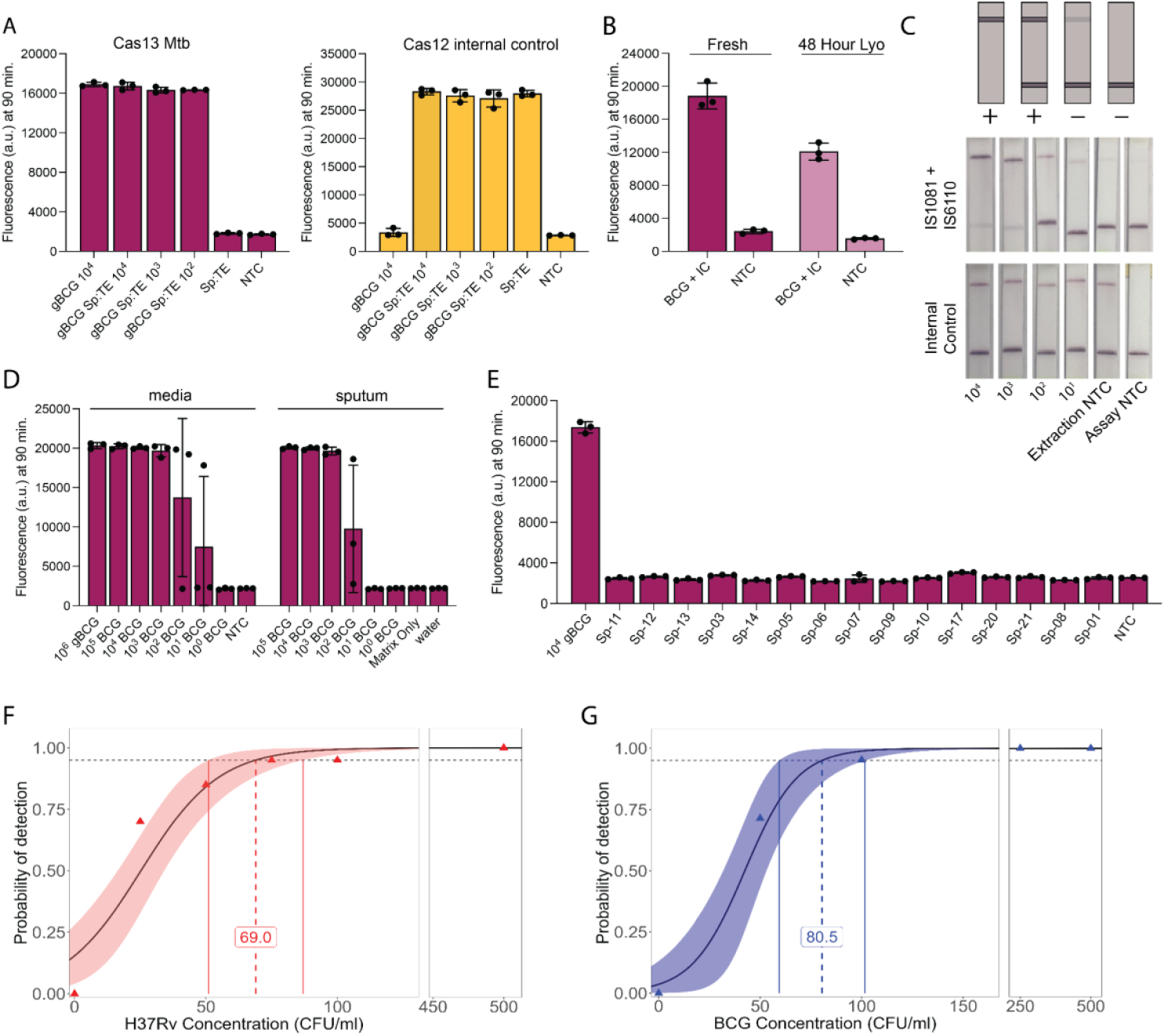
Validation of Cas13a dual detection assay against clinically relevant samples. A. Cas13a detecting BCG genomic DNA spiked into sputum:TE matrix and Cas12a detecting internal control. Sp is sputum and gBCG is genomic BCG. Units are genomes/µL. Cas13a reporter is FAM and Cas12a reporter is HEX. **B**. Comparison of fresh Cas13a (left) or lyophilizer Cas13a (right) assays detecting synthetic internal control (IC) and BCG genome. Synthetic targets were mixed at a concentration of 10^4^ and 10^3^ copies/µL, respectively. **C**. Lateral Flow read-out of Cas13a detecting dual targets IS6110+IS1081 (top) and Cas12a detecting internal control (bottom) in BCG spiked into sputum:TE matrix. Appearance of the top line is a positive readout. Faint lines are interpreted as negative. Disappearance of the bottom line is not necessary. **D**. Range finding experiment demonstrating the dynamic range of fresh Cas13a dual detection assay. Cas13a detecting BCG and Cas12a detecting internal control. BCG culture (left) and BCG spiked into sputum:TE (right). gBCG is genomic BCG and units are genomes/µL. All other units are CFU/mL. **E**. Testing fresh Cas13a and Cas12a coupled assay against TB-negative clinical sputum samples. Numbers indicate patient number. **F**. Limit of detection (LoD) for H37Rv. LOD = 69.0 (51.0 – 86.9). **G**. Limit of detection (LoD) for BCG. LOD = 80.5 (59.4 – 101.6). In A, B, D, and E, error bars: SD based on n=3. C shows a single representative sample. F (N=20) and G (N=21), was plotted using ggplot2 in R.

For ease of use, we attempted to combine Cas12a and the dual detection Cas13a assay in a single SHINE reaction. However, this integration reduced sensitivity and slowed the rate of signal accumulation for both enzymes (Supplementary Fig. 6). We found that excluding any one of the three amplification-detection sets restored this sensitivity, likely the result of competing exponential demands on shared RPA machinery (Fig. 2 and Supplementary Fig. 6a). As sensitivity and sample coverage are critical to the utility of any diagnostic assay, we opted to split the Cas12a internal control and Cas13a dual detection assay into two chambers, one with the internal control and one dedicated to *Mtb* detection. As these assays are run from the same processed sample and performed in parallel, the separation of components should minimally increase labor. Herein, this parallelised assay format is referred to as SHINE-TB.

As TB cases are most prevalent in low- and middle-income countries (LMICs), it is critical to develop accessible and field deployable assay format. Similar molecular diagnostics have previously been shown to be amenable to freeze-drying with minimal effects on assay performance^9,13^. We lyophilized single-use tubes of SHINE-TB for 24 hours and stored them at 4°C for 48 hours before testing the resuspended assays on BCG-spiked sputum, yielding comparable results to their fresh assay counterparts, albeit with a reduction to maximum fluorescence in this fluorometric assay format (Fig. 3b and Supplementary Fig. 5b and 5c). We also converted the reporter to be compatible with a lateral flow readout, whereby the quencher at one end of the reporter is replaced by biotin; intact reporters are immobilized at a lower streptavidin band while the antigenic ends of cleaved reporters freely migrate past to reach the upper band (disappearance of the lower biotin line is not required; faint positive test lines are interpreted as negative; Fig. 3c and Supplementary Fig. 7). The assay consistently detected the internal control and 10^2^ CFU/mL of BCG like its fluorometric counterpart.

### Characterizing Assay Performance in Contrived Clinical Samples

To determine the dynamic range of the final fluorometric SHINE-TB assay, we tested pooled sputum samples spiked with BCG cultures at various concentrations. BCG culture was used as positive control. We found that sputum compared to media did not change the upper limit of detection but did affect the lower limit of the Cas13a dual detection assay, where the samples at 10^2^ CFU/mL (colony-forming units (CFU)/mL BCG in pooled samples (representing between 0 and 5 copies of *IS6110* per assay, based on digital PCR (dPCR)) appeared to be near the threshold of detection (Fig. 3d, Supplementary Tables 1 and 2, and Supplementary Fig. 8a). Internal control signals remained unchanged irrespective of background BCG concentrations (Supplementary Fig. 8b and c).

We then tested the specificity of fresh SHINE-TB against individual sputum samples from fourteen TB-negative patients to mimic real-world sample variability. Sputum samples were collected from TB unsuspected patients with respiratory symptoms and were confirmed *Mtb* negative using dPCR (Fig. 3e). The *Mtb*-negative samples did not elicit any false positive assay detections, although two samples, Sp-11 and Sp-09, tested negative for the internal control, suggesting variable sample extraction or assay inhibition (Supplementary Fig. 8d). Of note, Sp-10 was a separate sputum sample collected from the same donor as Sp-09, which did not inhibit the assay, showing that there is variability even in samples collected from the same patient.

The Limit of detection (LoD) of the Cas13a dual detection assay was established with various concentrations of BCG (50-750 CFU/mL) and H37Rv (25-500 CFU/mL) spiked into confirmed *Mtb* negative pooled sputum; DNA was extracted using the sample processing described in the methods^31^. LoD fluorescence cutoffs were established as the average fluorescence of the sputum-only controls plus three times their standard deviation (SD). The LoD was defined as the concentration at which this threshold was reached by 95% of the replicates at a single concentration, as inferred by a logistic regression curve. The LoD for BCG was determined to be 80.5 (n positive/N total, 59.4 – 101.6, 95% CI) CFU/mL (Fig. 3f and Supplementary Fig. 9a and e). All concentrations above 100 CFU/mL were detected, and 15/21 50 CFU/mL samples were detected. At 50 CFU/mL, there is ∼1 copy/µL of *IS6110* in the input sample, ∼2 copies per 20 µL reaction, according to dPCR (Supplementary Table 3). Note, due to the low copy number and high viscosity of sputum, samples containing below 500 CFU/mL required mixing by pipetting 10+ times. All samples were positive for the internal control (Supplementary Fig. 9b). The LoD for H37Rv was 69.0 (51.0 – 86.9, 95% CI) CFU/mL (Fig. 3g and Supplementary Fig. 9c, f, and g) and all samples were detected by the internal control assay (Supplementary Fig. 9d).

### SHINE-TB Performs Well on Patient Samples

We performed a clinical validation of SHINE-TB on fourteen sputum samples collected from a convenience sample of symptomatic adults undergoing evaluation for pulmonary tuberculosis in two public primary healthcare networks in Cali, Colombia. Median age was 44 years old (range 32 - 59), 28.6% (4/14) were female, and 7.1% (1/14) were HIV-positive with a mean cough duration of 30 days (range: 20 - 60). Samples were characterized by smear microscopy, MGIT (liquid) culture, and GeneXpert MTB/RIF Ultra (‘Ultra’). Among the 14 samples, 6 were MGIT culture-positive (all smear microscopy positive with low-high positive Ultra results), 7 were MGIT culture-negative, and 1 was contaminated (indeterminate culture status).

The samples were processed using our sample processing method described in the methods for use in SHINE-TB and dPCR assays (Fig. 4a). SHINE-TB identified all 6 culture-positive samples as positive (100% sensitivity) and all 7 of the culture-negative samples as negative (100% specificity) (Fig. 4b, Supplementary Fig. 10, and Supplementary Table 5). This compared favorably to Ultra, which showed 100% sensitivity (6/6) and 86% specificity (6/7; 1 Ultra low-positive, culture-negative sample) against the single MGIT culture (Fig. 4b, 4c, and Supplementary Table 4). The one sample with a contaminated/indeterminate culture was negative by smear, Ultra, and SHINE-TB. For further information on the samples, see Supplementary Table 4.

**Figure 4.**
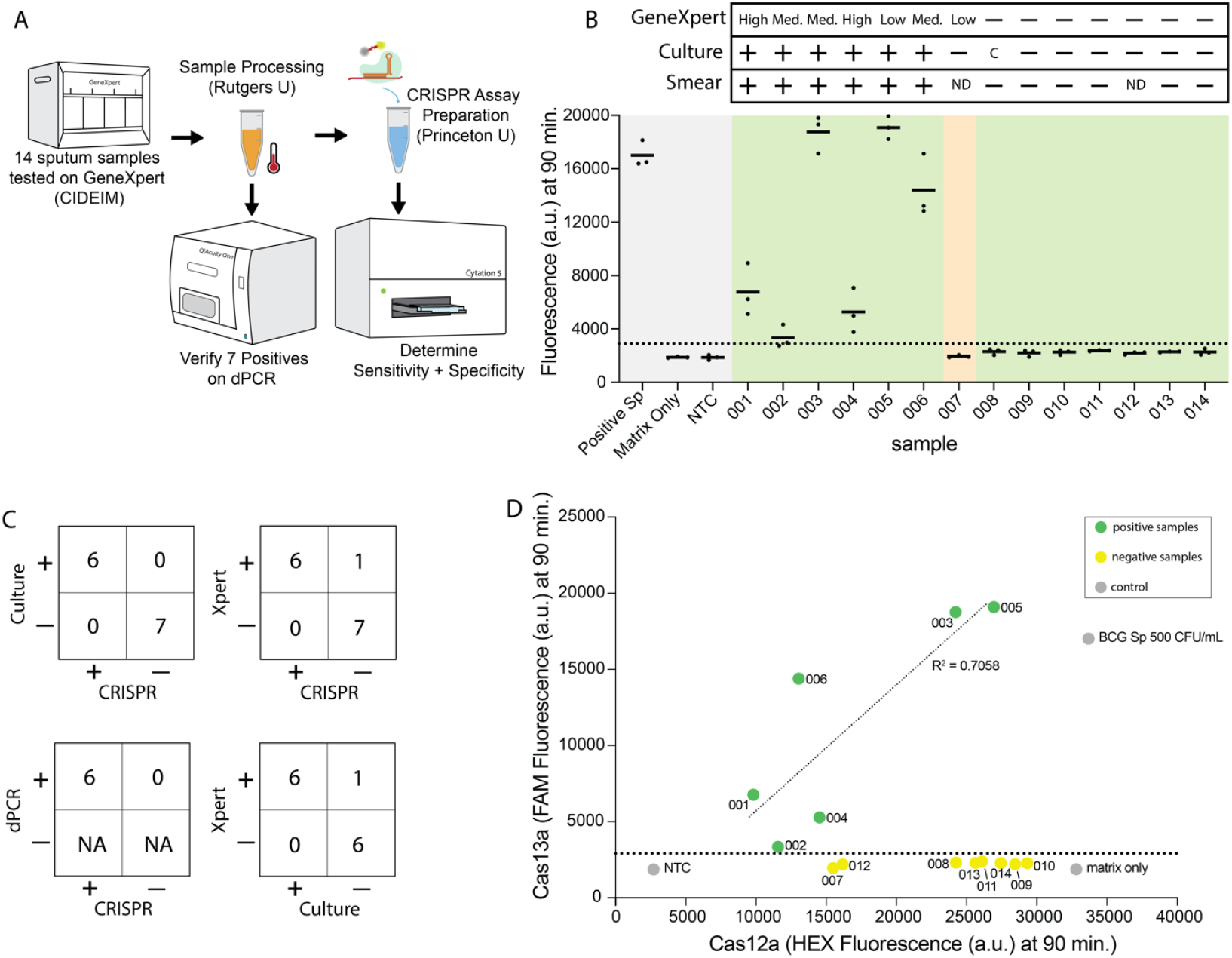
TB Detection from Sputum using Fresh SHINE-TB. A. Schematic showing workflow of patient sample testing. Sputum samples were collected and tested using GeneXpert at CIDEIM in Colombia. Samples were shipped to Rutgers for novel sample processing. Processed samples were used for CRISPR assays and dPCR. **B**. TB detection in 14 different sputum samples using Cas13a dual detection. For each sample, a line is shown at the mean of 3 technical replicates. Shading: Grey - Controls, Green - correct call, Yellow - unmatched call for GeneXpert vs CRISPR and culture. LoD is shown as a dotted line at 2900 a.u. Triplicates were averaged for determination of positive and negative. Positive sputum is 500 CFU/mL. Plus signs mean positive detection and minus signs mean no detection. C means contamination/not determined. ND means not done or data not collected. **C**. Tables showing Cas13a dual detection compared to culture (top left), GeneXpert (top right), and dPCR (middle right). Table showing GeneXpert vs Culture (bottom right). Further details in Supplementary Table 4. **D**. Average Cas13a (FAM) vs Cas12a internal control (HEX) signal for each sample seen in (B). Linear trend was determined for the positive samples with an R squared value of 0.7058.

We also plotted the signal intensity for our MTBC vs internal control assays (Fig. 4d). For the - samples with positive MTBC signal, there was a positive correlation between Cas13a signal and Cas12a signal, supporting that internal control performance reflects the performance of the MTBC assay and thus may help indicate issues with sample inhibition.

## Discussion

We have demonstrated the accuracy and feasibility of SHINE-TB for the detection of MTBC DNA and human DNA. We developed and compared two separate Cas12a- and Cas13a-based assays for the detection of MTBC, finding that Cas13a performs better for the purposes of single-pot, manipulation-free diagnostic assays. As high-burden TB countries often face significant resource limitations, isothermal techniques provide equipment free ways to amplify nucleic acids. Loop-mediated isothermal amplification (LAMP) is most widely used isothermal amplification method but requires incubation at 60°C-65°C for the duration of the reaction and is known to occasionally produce false positive results because it relies on numerous long primers that each have a non-negligible probability of off-target hybridization^36–42^. Recombinase polymerase amplification (RPA) can amplify sequences at 37°C using only a pair of primers per amplicon, but can be subject to non-specific amplification when longer primers are used for more efficient reactions, while the integration of CRISPR with RPA may help mitigate these issues with an added layer of specificity checking^14,22,30,43–46^.

Our assays were designed to create a one-step procedure that does not require a mechanical separation between isothermal amplification and detection. Despite integrating suboptimal PAM sequences to mitigate the competition between Cas12a and amplification^33^, we found that Cas13a had greater sensitivity than Cas12a. This demonstrates the benefits of an efficient workflow mitigating the complexity of longer primers to initiate transcription and additional substrates needed for T7 RNA polymerase. Cas13 has been favored in other studies as well, possibly due to its higher turnover for trans-cleavage^12,13,23,47,48^.

We then expanded this to a Cas13a dual detection assay for other strains in the MTBCs with varying *IS6110* and *IS1081* copy numbers^49–51^. Although we favored Cas13a for a one-pot test that required sensitivity and robustness, Cas12a exhibited appropriate sensitivity for an internal control targeting a highly abundant genetic element in properly extracted human samples, as standard samples had significant signal saturation.

We have demonstrated the feasibility of lyophilization and lateral flow readout to provide a means for deployability to lower resource areas. If the two were combined, and considering the robust sample preparation, this would allow for minimal sample handling with minimal equipment. Entirely equipment-free SHINE is possible as 37°C is the optimal temperature for both RPA and Cas13a^13^. Previous CRISPR-based TB studies have successfully incorporated lateral flow^23^ but not in a one-pot format. This is the first TB test where amplification and detection are combined that is compatible with both lateral flow and lyophilization.

Compared to GeneXpert MTB/RIF cartridges (1mL), SHINE-TB uses significantly less sputum sample volume (100 µL per extraction and ∼2 µL per reaction)^52^. Despite contributing just 5% of the overall reaction volume, some individual sputum types showed significant assay-inhibition for both Cas13a and Cas12a using the crude sample prep, possibly due to inhibition of RPA^53^. Additional experiments are required to fully understand the mechanisms of inhibition, however, if the internal control also fails, the test samples will be called “indeterminates”, requiring repeat testing.

Combined with the rapid sample processing and optimized Cas13a SHINE-TB assay for *Mtb* spiked in sputum, we found an LoD of ∼80 CFU/mL for H37Rv, which was slightly higher than the published LoD for Ultra (15.6 CFU/ml) and ∼60 CFU/mL for BCG, which was comparable or better than the published Ultra BCG LoD of 143.3 CFU/ml^54^. Considering the expected higher copy numbers of both IS6110 and IS1081 in *Mtb* H37Rv compared to M.bovis BCG, we expected the H37Rv LoD to be much better (Supplementary Fig. 9e - 9g). Since dPCR testing of 50 CFU/mL BCG detected ∼1 copy/µL of IS6110, the target genomes may follow a Poisson distribution, meaning some reactions may randomly contain no genomes at low concentrations (Supplementary Table 3). Thus, at these concentrations, increased copy number does not significantly increase detection. Regardless, SHINE detected *Mtb* in a small set of smear-positive clinical samples with 100% specificity and 100% sensitivity compared to culture. While this small clinical evaluation supports the promise of an initial SHINE-TB prototype, future dedicated clinical studies with more participants and participant diversity, as well as smear-negative, culture-positive samples are needed for more comprehensive and precise performance measures.

Additional steps must be taken to address broader needs for the field. As drug-resistant strains of *Mtb* become increasingly prevalent, many diagnostics include drug susceptibility testing^54,55^. Preferably, a diagnostic would be able to identify resistance for first-line drugs, such as rifampicin (Rif). Increased multiplexing is necessary to achieve this goal, as current amplification restrictions prevent us from amplifying more than two targets. In the future it would be useful to increase multiplexing capacity by changing amplification strategies or by spatially isolating parallel reactions, as previously done^11,47,56^. Additional improvements would expand the compatibility of the assay to multiple sample types, as sputum can be difficult for some patients to produce^57,58^. Ideally, our platform would be compatible with sample types such as saliva and oral swabs^59^. Further testing will be needed to determine which sample types are amenable to extraction and diagnosis, while better biochemical characterizations should help identify compounds that are inhibitory to the overall diagnostic process. Our simple one-step assay provides robust detection of both MTBC and human DNA, taking us one step closer to accessible and reliable detection of this deadly disease.

## Methods

### Synthetic targets and genomic DNA

Gene fragments for IS6110 and IS1081 were ordered from Integrated DNA Technologies (IDT) (sequences listed in supplementary files) and resuspended in nuclease-free water. Bacille Calmette-Guerin (BCG) genomic DNA was ordered from ATCC (catalog 35734D-2) and quantified with dPCR. Quantitative *Mtb* genomic DNA was ordered from ATCC (25177DQ).

### Sequence Design

Cas12a target sequences were chosen by first compiling canonical sequences and related diversity information for chosen insertion sequences or possible internal control genes using BLAST^60^ and Clustal Omega^61^ on publicly available datasets. We then annotated these sequences for all canonical and permitted non-canonical Cas12a PAM sequences and ranked candidates manually for conservation throughout the amplified region and the energetics of crRNA ensemble conformations^62^. Cas13a designs were generated using ADAPT ^34^on the same consensus sequences with 40 nt primers and otherwise default parameters, before being manually ranked for conservation and free energy, as above. The Cas13a crRNA IS6110 C had a slightly lower ADAPT rating than the other chosen candidates but was made to overlap with Cas12a cr5 to compare the two assays’ efficiencies more directly. A T7 RNA polymerase promoter was added to the 5’ end of all forward primers for Cas13a assays. All crRNAs, coupled primers, and synthetic targets used in this publication are listed in the supplement.

### Final sequences used in this study are mentioned in supplementary tables 6 and 7. Two-step and one-step Cas12a assays

Two-step Cas12a assays were performed by mixing an amplification master mix (below) with target, allowing amplification to occur for 30 minutes before adding Cas12-crRNA RNPs (Figure 1B and 1C). Master mixes (concentrations given for final reaction) were generated by combining SHINE buffer (20 mM HEPES pH 8.0, 60 mM KCl, 3.5% PEG-8000, nuclease-free (nf) water), 1 *U*/µL RNAse Inhibitor (New England BioLabs), 70 nM of each RPA primer, 0.25 µM FAM 5C quenched reporter (IDT), and 14 nM MgOAc in nf water. TwistAmp Basic RPA pellets were used for amplification, whereby 1 pellet was resuspended for every 107.5 µL of final reaction volume. The target (10% of final reaction volume) and amplification master mix were combined, lightly vortexed, and incubated at 37 °C for 30 minutes in an Agilent BioTek Cytation 5 microplate reader, taking fluorescence readings every 5 minutes (excitation: 485, emission 525). crRNAs and *Lb*Cas12a Ultra (10007922; IDT) in nf water at 10X their final equimolar concentration of 4 or 20 nM were combined on ice for 15-20 minutes and added to each reaction 30 minutes into the reaction. 15 µL reactions were performed in technical triplicates (except for two early optimization experiments performed in duplicate to increase condition throughput) in 384-well clear-bottom microplates (788096; Greiner). Reactions were then returned to the plate reader at 37 °C for up to 3 hours of additional fluorescence readings.

Single-step Cas12a assays were performed by mixing complete Cas12a master mix (90% by volume) with target (10% by volume) (Fig. 1c, 1d, 1e, 1f, and 2d). Master mixes were prepared as in the two-step protocol, except crRNAs and Cas12a were added in the master mix before adding target, for a final concentration of 4 nM (Fig. 1 and 2), 30 nM (Fig. 3), or as otherwise indicated. Corning 384-well microplates (CLS3544) were used for 15 µL reactions (Fig. 1, 2, and 3). Reactions were incubated and measured as above.

### Single-step Cas13a assays

Single-step Cas13a assays were performed as for the single-step Cas12a reactions, except that to form the Cas13a master mix, the same SHINE buffer and 14 nM MgOAc were mixed with 45 nM *Lwa*Cas13a (Genscript, stored in 100 mM Tris HCl pH 7.5 and 1 mM DTT), 0.3 nM of each rNTP (New England Biolabs), 1 *U*/mL T7 RNAP (Biosearch Technologies), 62.5 nM FAM 6U quenched reporter (5’-/56-FAM/rUrUrUrUrUrU/3IABkFQ/-3’; IDT), 45 nM of a single crRNA (Fig. 2a, 2b, and 2c) or 22.5 nM of each crRNA in a combination (Figure 2c, 2d, Fig. 3, and Fig. 4), and 70 nM of the appropriate RPA primers in nf water. Reactions were loaded in technical triplicates into Greiner 384-well clear-bottom microplates for 15 µL reactions (Fig. 2D) or Corning 384-well microplates for 20 µL reactions (Fig. 3 and 4). The reaction was then incubated at 37 °C for up to 3 hours in a Cytation 5, as above. Single-step Cas13a assays were performed by mixing complete Cas12a master mix (90% by volume) with target (10% by volume). When the target is sputum, pipette mix 10+ times and pulse vortex before spinning down.

Combined Cas12a/Cas13a assays were performed by combining all the non-redundant reagents from the two independent single-pot assays and replacing the DNA reporter with 0.25 µM HEX 5C quenched reporter (IDT), to be measured in two fluorescence channels: excitation: 485, emission 525; excitation: 530, emission: 570.

### Lateral flow read-outs

The assay was adapted to lateral flow with the following conditions: the quenched Cas13a reporter was replaced with 750 nM FAM 14U Bio (IDT) and the Cas12a reporter was replaced with FAM 5C Bio (IDT) when alone or FAM 5C Dig (IDT). Reactions were incubated in PCR strip tubes at 37 °C for 90 minutes before being diluted 1:4 in HybriDetect Assay buffer (Milenia). The reaction was equilibrated for 5 minutes before adding HybriDetect lateral flow strips for 1 minute. Images were taken 1 minute after dipstick removal.

### Lyophilization

Lyophilization methods were based on a previously described protocol^13^, with some minor modifications. The dual assay was optimized to ensure integrity after lyophilization. The reaction was prepared as described above, except 20 mM HEPES (pH 8.0) was added in place of the complete SHINE buffer. Sucrose at 5% (w/v) and mannitol at 150 mM, were added as cryoprotectants. Assays were aliquoted (eight reactions per aliquot), flash frozen, and lyophilized at -30 °C for 24 hours using a Freezone Triad Benchtop Freeze Dryer from Labconco. Aliquots were then vacuum sealed alongside a desiccant and stored in 4 °C until further use. Assays were resuspended in 3.5% PEG, 60 mM KCl, and 14 mM MgOAc in nf water, aliquoted, and mixed with target. Reactions were loaded in technical triplicates in Corning 384-well microplates for 20 µL reactions (Fig. 3). Reactions were then incubated at 37 °C for up to 3 hours. Fluorescent measurements were taken every 5 minutes in a Cytation 5 microplate reader, as above.

### Collection of Clinical Samples for Contrived Sample Evaluations

TB-negative samples for spiking were collected from patients of University Hospital, Newark New Jersey, USA, after written informed consent (Rutgers eIRB #Pro2020001138). Eligible participants were adults > 18 years old diagnosed with a non-TB respiratory condition (e.g. cardiogenic pulmonary edema, asthma exacerbation, other bacterial pneumonia) and were willing and able to provide an expectorated sputum sample.

Following written informed consent, symptomatic individuals undergoing evaluation for tuberculosis (TB) at two primary health centers in Cali, Colombia, provided expectorated sputum for examination. The study protocol was reviewed and approved by the Institutional Review Boards (IRBs) for ethical conduct in research involving human subjects at the Centro Internacional de Entrenamiento e Investigaciones Médicas (CIDEIM # 1325). The protocol adhered to both national regulations (Resolution 008430, Ministry of Health, Republic of Colombia, 1993) and international ethical guidelines, including the Declaration of Helsinki and its amendments (World Medical Association, Fortaleza, Brazil, October 2013).

### Sample Processing

Sample processing was performed as previously described^31^. Briefly: Chelex extraction for sputum samples: For individual patient screening, inhibitory patient screening, and dynamic range extractions. 100 µL of spiked sputum was aliquoted into a screw-cap Lysing Matrix B tube (MP Biomedicals, Cat. #MP116911100), subsequently 200 µL of Chelex100 resin (instagene Matrix BIO-RAD, Cat. #7326030) was added. Samples were vortexed three times, for 30 seconds at 3400 RPM each time, subsequently incubated at 95C for 30 minutes, then centrifuged at 9400 RCF for 2 minutes. ∼60-80 µL of supernatant were collected to serve as inputs for the diagnostic assays.

LoD Extractions: 400 µL of spiked sputum was aliquoted into a screw-cap Lysing Matrix B tube, subsequently 800 µL of Chelex100 resin (instagene Matrix) was added. Samples were vortexed three times, for 30 seconds at 3400 RPM each time, subsequently incubated at 95C for 30 minutes, then centrifuged at 9400 RCF for 2 minutes. ∼800 µL of supernatant was collected.

A convenience sample of symptomatic adults aged ≥18 years and undergoing evaluation for tuberculosis (TB) at two primary health centers in Cali, Colombia, provided two expectorated or induced sputum samples on the same day. One was sent for smear microscopy and mycobacterial culture on liquid media (MGIT) on-site in the clinical laboratories, while the other was sent for molecular testing using the GeneXpert MTB/RIF Ultra assay at the CIDEIM research laboratory in accordance with the manufacturer’s instructions. Participants were selected the following way: Research staff (a nursing assistant or a physician) approached consecutive individuals referred for TB microbiological testing according to the laboratory electronic records from district hospitals located in the central and eastern parts of the city (ESE Centro and ESE Oriente). In addition, the research staff screened a convenience sample of participants in the emergency, respiratory therapy, and outpatient departments for TB symptoms, and encouraged clinicians to refer them for TB diagnostic testing.

### Spiked and Contrived Sample Testing

Clinical samples were prepared using methods described above. Primers, crRNAs, and master mix are the same as above.

dPCR was performed in the QiAcuity digital PCR system using QiAcuity 4X Probe PCR kit and 26k 8-well nanoplate (QIAGEN). Previously published sequences for primers and probes were used (IS6110-I-F, IS6110-I-R, and IS6110-TM)^54^. For the final reaction volume of 12µl, 3µl of 4X Probe PCR Master mix, 0.8µM concentration of forward and reverse primers, 0.4µM concentration of probe, and 2µl of DNA sample were added. Molecular grade, nuclease free water was used to complete the volume. Thermal cycling conditions are: PCR initial heat activation at 95°C for 2 minutes, followed by 40 cycles of denaturation at 95°C for 15 seconds and combined annealing/extension at 60°C for 30 seconds. DNA samples were tested both directly and at 10-fold dilution in replicates of three. M. *bovis* BCG DNA at 100gc/reaction, and nuclease free water were used as positive and negative control respectively.

### Data analysis

#### Plots and graphs were generated using Prism software

Data were plotted in R using the ggplot2 package. Simple logistic regression was used to model detection probabilities for (a) H37RV and (b) BCG bacteria as a function of concentration (Figure 3F). The 95% LOD was estimated as the concentration in which the probability of detection of a positive sample is 0.95. 95% Wald confidence intervals were generated from the standard error of the estimated concentrations.

Assay cutoffs for positive or negative results were derived using a 70:30 train-test split strategy of spiked sputum data with known bacterial concentrations ranging from 0 (Mtb-negative samples) to 10^4^ cfu/mL (Mtb-positive samples). ROC curves were constructed from a dataset of 115 experiments with known bacterial concentrations. Four approaches were explored for setting the cutoff; (1) the mean of the negative samples plus three times the standard deviation of the negative samples, (2) the point on the ROC curve corresponding to the maximum Youden index, (3) the point on the ROC curve corresponding to the WHO 2024 Target Product Profile (TPP) sensitivity standard for a sputum-based near-point-of-care test and (4) the point on the ROC curve corresponding to the WHO 2024 specificity standard for all diagnostic tests (Supplementary Table 5). Both the Youden index and the specificity standard approaches gave a cutoff of 2895.42 fluorescent units, which was selected and rounded to 2900 for the final cutoff. This cutoff was then applied to evaluating SHINE performance among the 14 clinical samples. The exact method for binomial confidence intervals was used to compute 95% confidence intervals for sensitivity and specificity estimates.

## Data Availability

All data produced in the present study are available upon reasonable request to the authors.

## Acknowledgments

We wish to thank the study participants, leaders and staff of the participating health facilities, and staff of the Centro Internacional de Entrenamiento e Investigaciones Médicas (CIDEIM) for their contributions to the clinical aspects of this study, and Lesly Fabiola Suarez for her help and support with securing patient samples. We would like to thank the Prud’homme Lab for allowing us to use their lyophilizer. Funding was provided by the National Institutes of Health (NIH) R21 AI174129 (J.L.D., D.A.V.), NIH R21 AI168808 (Y.L.X., P.B., C.M.), NIH R01 AI182281(Y.L.X, C.M.), New Jersey Alliance for Clinical and Translational Science (NJACTS) UL1TR003017 (Y.L.X., P.B., C.M.) and Centers for Disease Control and Prevention 75D30122C15113 (C.M.). A.G.B. was supported by NIH Training Grants T32GM148739 and T32GM007388.

## Author Contributions

O.R.S.D., A.G.B., N.M., P.B., Y.L.X, and C.M. designed research. A.G.B., O.R.S.D., N.M., Y.H., and S.T. performed experiments. A.G.B, O.R.S.D, and R.R. performed data analysis. A.G.B., O.R.S.D., and C.M. wrote the article with input from all coauthors.

## References

1. World Health Organization. Global Tuberculosis Report 2024. WHO. https://www.who.int/teams/global-tuberculosis-programme/tb-reports/global-tuberculosis-report-2024 (2024).

2. Demers, A.-M. et al. High yield of culture-based diagnosis in a TB-endemic setting. BMC Infect. Dis. 12, 218 (2012).

3. Kunkel, A. et al. Smear positivity in paediatric and adult tuberculosis: systematic review and meta-analysis. BMC Infect. Dis. 16, 282 (2016).

4. Jenkins, H. E. et al. Incidence of multidrug-resistant tuberculosis disease in children: systematic review and global estimates. Lancet 383, 1572–1579 (2014).

5. Omar, M., el Naggar, M., AboYoussef, S. & Eltanty, A. Assessment of the participation of primary care services in national tuberculosis control program in El Behaira Governorate. Egypt. J. Chest Dis. Tuberc. 69, 289–295 (2020).

6. Hsiang, E. et al. Higher cost of implementing Xpert(®) MTB/RIF in Ugandan peripheral settings: implications for cost-effectiveness. Int. J. Tuberc. Lung Dis. 20, 1212–1218 (2016).

7. Boehme, C. C. et al. Feasibility, diagnostic accuracy, and effectiveness of decentralised use of the Xpert MTB/RIF test for diagnosis of tuberculosis and multidrug resistance: a multicentre implementation study. Lancet 377, 1495–1505 (2011).

8. Schumacher, S. G. et al. Guidance for Studies Evaluating the Accuracy of Sputum-Based Tests to Diagnose Tuberculosis. J. Infect. Dis. 220, S99–S107 (2019).

9. Gootenberg, J. S. et al. Nucleic acid detection with CRISPR-Cas13a/C2c2. Science 356, 438–442 (2017).

10. Myhrvold, C. et al. Field-deployable viral diagnostics using CRISPR-Cas13. Science 360, 444–448 (2018).

11. Ackerman, C. M. et al. Massively multiplexed nucleic acid detection with Cas13. Nature 582, 277–282 (2020).

12. Arizti-Sanz, J. et al. Streamlined inactivation, amplification, and Cas13-based detection of SARS-CoV-2. Nat. Commun. 11, 5921 (2020).

13. Arizti-Sanz, J. et al. Simplified Cas13-based assays for the fast identification of SARS-CoV-2 and its variants. Nat Biomed Eng 6, 932–943 (2022).

14. Chen, J. S. et al. CRISPR-Cas12a target binding unleashes indiscriminate single-stranded DNase activity. Science 360, 436–439 (2018).

15. Li, S.-Y. et al. CRISPR-Cas12a has both cis- and trans-cleavage activities on single-stranded DNA. Cell Res. 28, 491–493 (2018).

16. East-Seletsky, A. et al. Two distinct RNase activities of CRISPR-C2c2 enable guide-RNA processing and RNA detection. Nature 538, 270–273 (2016).

17. Abudayyeh, O. O. et al. C2c2 is a single-component programmable RNA-guided RNA-targeting CRISPR effector. Science 353, aaf5573 (2016).

18. Tambe, A., East-Seletsky, A., Knott, G. J., Doudna, J. A. & O’Connell, M. R. RNA Binding and HEPN-Nuclease Activation Are Decoupled in CRISPR-Cas13a. Cell Rep. 24, 1025–1036 (2018).

19. Gootenberg, J. S. et al. Multiplexed and portable nucleic acid detection platform with Cas13, Cas12a, and Csm6. Science 360, 439–444 (2018).

20. Feng, W., Peng, H., Zhang, H., Weinfeld, M. & Le, X. C. A Sensitive Technique Unravels the Kinetics of Activation and Trans-Cleavage of CRISPR-Cas Systems. Angew. Chem. Int. Ed Engl. 63, e202404069 (2024).

21. Nalefski, E. A. et al. Kinetic analysis of Cas12a and Cas13a RNA-Guided nucleases for development of improved CRISPR-Based diagnostics. iScience 24, 102996 (2021).

22. Peng, L. et al. Rapid detection of Mycobacterium tuberculosis in sputum using CRISPR-Cas12b combined with cross-priming amplification in a single reaction. J. Clin. Microbiol. 62, e0092323 (2024).

23. Thakku, S. G. et al. Genome-wide tiled detection of circulating Mycobacterium tuberculosis cell-free DNA using Cas13. Nat. Commun. 14, 1803 (2023).

24. Jia, N. et al. A CRISPR-Cas12a-based platform for ultrasensitive rapid highly specific detection of Mycobacterium tuberculosis in clinical application. Front. Cell. Infect. Microbiol. 13, 1192134 (2023).

25. Sam, I. K. et al. TB-QUICK: CRISPR-Cas12b-assisted rapid and sensitive detection of Mycobacterium tuberculosis. J. Infect. 83, 54–60 (2021).

26. Huang, Z. et al. CRISPR detection of circulating cell-free Mycobacterium tuberculosis DNA in adults and children, including children with HIV: a molecular diagnostics study. Lancet Microbe 3, e482–e492 (2022).

27. Ai, J.-W. et al. CRISPR-based rapid and ultra-sensitive diagnostic test for Mycobacterium tuberculosis. Emerg. Microbes Infect. 8, 1361–1369 (2019).

28. Wang, Y. et al. LAMP-CRISPR-Cas12-based diagnostic platform for detection of Mycobacterium tuberculosis complex using real-time fluorescence or lateral flow test. Mikrochim. Acta 188, 347 (2021).

29. Peng, L., Fang, T., Dai, L. & Cai, L. Diagnostic value of cross-priming amplification combined with CRISPR-Cas12b in detecting cell-free DNA in tuberculous pleural effusion. Open Forum Infect. Dis. 11, ofae674 (2024).

30. Lobato, I. M. & O’Sullivan, C. K. Recombinase polymerase amplification: Basics, applications and recent advances. Trends Analyt. Chem. 98, 19–35 (2018).

31. Modi, N. et al. Simplified Co-extraction of total Nucleic Acids from Respiratory Samples for detection of Mycobacterium tuberculosis and SARS-CoV-2 optimized for compatibility across Diagnostic Platforms. medRxiv 2025.02.27.25322880 (2025) doi:10.1101/2025.02.27.25322880.

32. Qiu, M., Zhou, X.-M. & Liu, L. Improved Strategies for CRISPR-Cas12-based Nucleic Acids Detection. J Anal Test 6, 44–52 (2022).

33. Lu, S. et al. Fast and sensitive detection of SARS-CoV-2 RNA using suboptimal protospacer adjacent motifs for Cas12a. Nat Biomed Eng 6, 286–297 (2022).

34. Metsky, H. C. et al. Designing sensitive viral diagnostics with machine learning. Nat. Biotechnol. 40, 1123–1131 (2022).

35. Lander, E. S. et al. Initial sequencing and analysis of the human genome. Nature 409, 860–921 (2001).

36. Nagai, K. et al. Diagnostic test accuracy of loop-mediated isothermal amplification assay for Mycobacterium tuberculosis: systematic review and meta-analysis. Sci. Rep. 6, 39090 (2016).

37. Iwamoto, T., Sonobe, T. & Hayashi, K. Loop-mediated isothermal amplification for direct detection of Mycobacterium tuberculosis complex, M. avium, and M. intracellulare in sputum samples. J. Clin. Microbiol. 41, 2616–2622 (2003).

38. Boehme, C. C. et al. Operational feasibility of using loop-mediated isothermal amplification for diagnosis of pulmonary tuberculosis in microscopy centers of developing countries. J. Clin. Microbiol. 45, 1936–1940 (2007).

39. Notomi, T. et al. Loop-mediated isothermal amplification of DNA. Nucleic Acids Res. 28, E63 (2000).

40. Howard, S. T., Oughton, M. T., Haddad, A. & Johnson, W. M. Absence of the genetic marker IS6110 from a strain of Mycobacterium tuberculosis isolated in Ontario. Can. J. Infect. Dis. 9, 48–53 (1998).

41. Gagneux, S. et al. Variable host-pathogen compatibility in Mycobacterium tuberculosis. Proc. Natl. Acad. Sci. U. S. A. 103, 2869–2873 (2006).

42. Meagher, R. J., Priye, A., Light, Y. K., Huang, C. & Wang, E. Impact of primer dimers and self-amplifying hairpins on reverse transcription loop-mediated isothermal amplification detection of viral RNA. Analyst 143, 1924–1933 (2018).

43. Singpanomchai, N. et al. Naked eye detection of the Mycobacterium tuberculosis complex by recombinase polymerase amplification-SYBR green I assays. J. Clin. Lab. Anal. 33, e22655 (2019).

44. Singpanomchai, N. et al. Rapid detection of multidrug-resistant tuberculosis based on allele-specific recombinase polymerase amplification and colorimetric detection. PLoS One 16, e0253235 (2021).

45. Boyle, D. S. et al. Rapid detection of Mycobacterium tuberculosis by recombinase polymerase amplification. PLoS One 9, e103091 (2014).

46. Mota, D. S. et al. Recombinase polymerase amplification in the molecular diagnosis of microbiological targets and its applications. Can. J. Microbiol. 68, 383–402 (2022).

47. Welch, N. L. et al. Multiplexed CRISPR-based microfluidic platform for clinical testing of respiratory viruses and identification of SARS-CoV-2 variants. Nat. Med. 28, 1083–1094 (2022).

48. Huyke, D. A. et al. Enzyme kinetics and detector sensitivity determine limits of detection of amplification-free CRISPR-Cas12 and CRISPR-Cas13 diagnostics. Anal. Chem. 94, 9826–9834 (2022).

49. Sreevatsan, S. et al. Restricted structural gene polymorphism in the Mycobacterium tuberculosis complex indicates evolutionarily recent global dissemination. Proc. Natl. Acad. Sci. U. S. A. 94, 9869–9874 (1997).

50. Cole, S. T. et al. Deciphering the biology of Mycobacterium tuberculosis from the complete genome sequence. Nature 393, 537–544 (1998).

51. Philipp, W. J. et al. An integrated map of the genome of the tubercle bacillus, Mycobacterium tuberculosis H37Rv, and comparison with Mycobacterium leprae. Proc. Natl. Acad. Sci. U. S. A. 93, 3132–3137 (1996).

52. Helb, D. et al. Rapid detection of Mycobacterium tuberculosis and rifampin resistance by use of on-demand, near-patient technology. J. Clin. Microbiol. 48, 229–237 (2010).

53. Schrader, C., Schielke, A., Ellerbroek, L. & Johne, R. PCR inhibitors - occurrence, properties and removal. J. Appl. Microbiol. 113, 1014–1026 (2012).

54. Chakravorty, S. et al. The New Xpert MTB/RIF Ultra: Improving Detection of Mycobacterium tuberculosis and Resistance to Rifampin in an Assay Suitable for Point-of-Care Testing. MBio 8, (2017).

55. Singh, V. & Chibale, K. Strategies to Combat Multi-Drug Resistance in Tuberculosis. Acc. Chem. Res. 54, 2361–2376 (2021).

56. Xu, Z. et al. Microfluidic space coding for multiplexed nucleic acid detection via CRISPR-Cas12a and recombinase polymerase amplification. Nat. Commun. 13, 6480 (2022).

57. Byanyima, P. et al. Feasibility and Sensitivity of Saliva GeneXpert MTB/RIF Ultra for Tuberculosis Diagnosis in Adults in Uganda. Microbiol Spectr 10, e0086022 (2022).

58. Luabeya, A. K. et al. Noninvasive Detection of Tuberculosis by Oral Swab Analysis. J. Clin. Microbiol. 57, (2019).

59. Nicol, M. P. et al. Microbiological diagnosis of pulmonary tuberculosis in children by oral swab polymerase chain reaction. Sci. Rep. 9, 10789 (2019).

60. Altschul, S. F., Gish, W., Miller, W., Myers, E. W. & Lipman, D. J. Basic local alignment search tool. J. Mol. Biol. 215, 403–410 (1990).

61. Madeira, F. et al. The EMBL-EBI Job Dispatcher sequence analysis tools framework in 2024. Nucleic Acids Res. 52, W521–W525 (2024).

62. Zadeh, J. N. et al. NUPACK: Analysis and design of nucleic acid systems. J. Comput. Chem. 32, 170–173 (2011).

