## Supplementary Information for "A Streamlined Point-of-Care CRISPR Test for Tuberculosis Detection Directly from Sputum"

**Supplementary Table 1.** dPCR data on sputum dilutions for Dynamic Range experiment 3D.

| (Sputum+BCG):TE IS6110 copies/µL | | | | | | |
|---|---|---|---|---|---|---|
| **CFU/mL** | **rep 1** | | **rep 2** | | **rep 3** | |
| **$10^5$** | 844.8 | 834.6 | 1048.8 | 1111.2 | 652.2 | 636.6 |
| **$10^4$** | 100.2 | 66.12 | 55.572 | 74.7 | 155.28 | 143.52 |
| **$10^3$** | 11.334 | 18.522 | 14.196 | 9.348 | 14.292 | 14.496 |
| **$10^2$** | 4.548 | 0 | 9.432 | N/A | 4.764 | 0 |
| **$10^1$** | 0 | 0 | 0 | 2.382 | 0 | 0 |
| **$10^0$** | 0 | 0 | 0 | 0 | 0 | 0 |
| **0** | 0 | 0 | 0 | 0 | 0 | 0 |
| **NTC** | 0 | 0 | | | | |

**Supplementary Table 2.** dPCR data on media dilutions for Dynamic Range experiment 3D

| (media+BCG):TE IS6110 copies/μL | | | | | | |
|---|---|---|---|---|---|---|
| **CFU/mL** | **rep 1** | | **rep 2** | | **rep 3** | |
| **$10^5$** | 1881.6 | 1873.2 | 2004 | 2146.2 | 1392 | 1381.2 |
| **$10^4$** | 249.18 | 264.72 | 119.34 | 112.68 | 81.42 | 129.48 |
| **$10^3$** | 14.37 | 7.182 | 18.708 | N/A | 23.028 | 15.978 |
| **$10^2$** | 0 | 2.406 | 2.424 | 0 | 5.028 | 0 |
| **$10^1$** | 0 | 0 | 0 | 2.4 | 0 | 0 |
| **$10^0$** | 0 | 0 | 0 | 0 | 0 | 0 |
| **0** | 0 | 0 | 0 | 0 | 0 | 0 |
| **NTC** | 0 | 0 | | | | |

.

**Supplementary Table 3.** dPCR results for BCG LoD.

| CFU/mL | Sputum:TE IS6110 copy # per uL | Media:TE IS6110 copy # per uL | Control |
|---|---|---|---|
| 1000 | 27.2 | 33.46 | NA |
| 750 | 28.44 | 43.12 | NA |
| 500 | 12.78 | 13.16 | NA |
| 250 | 9.64 | 11.1 | NA |
| 100 | 3.22 | 4.42 | NA |
| 50 | 1.08 | 1.1 | NA |
| Negative Control | 0 | 0 | NA |
| H37Ra gDNA $10^3$ copies | NA | NA | 44238 |
| NTC | NA | NA | 0 |

**Supplementary Table 4.** Patient sample information for Fig. 4.

| # | Patient # | Xpert (Colombia) | Xpert Ct Value (Colombia) IS-1081/6110 | dPCR copies/2µL (Rutgers) IS-1081/6110 | Description |
|---|---|---|---|---|---|
| 001 | TB-02-0019 | Medium | 16.2 | 10890.06667 | Smear (+), Culture (+), GeneXpert sputum (+) |
| 002 | TB-01-0083 | High | 15.9 | 16065.7 | Smear (+), Culture (+), GeneXpert sputum (+) |
| 003 | TB-01-0111 | Medium | 15.8 | 358.6633333 | Smear (+), Culture (+), GeneXpert sputum (+) |
| 004 | TB-01-0112 | High | 15.9 | 37198 | Smear (+), Culture (+), GeneXpert sputum (+) |
| 005 | TB-01-0120 | Low | 16.5 | 34 | Smear (+), Culture (+), GeneXpert sputum (+) |
| 006 | TB-01-0159 | Medium | 16 | 3453.76 | Smear (+), Culture (+), GeneXpert sputum (+) |
| 007 | TB-01-0162 | Low | 20.2 | 0 | Smear (ND), Culture (-), GeneXpert sputum (+) |
| 008 | TB-02-0018 | Not Detected | 0 | NA | Smear (-), Culture (C), GeneXpert sputum (-) |
| 009 | TB-02-0020 | Not Detected | 0 | NA | Smear (-), Culture (-), GeneXpert sputum (-) |
| 010 | TB-02-0021 | Not Detected | 0 | NA | Smear (-), Culture (-), GeneXpert sputum (-) |
| 011 | TB-02-0022 | Not Detected | 0 | NA | Smear (-), Culture (-), GeneXpert sputum (-) |

| | | | | | |
|---|---|---|---|---|---|
| 012 | TB-02-0023 | Not Detected | 0 | NA | Smear (ND), Culture (-), GeneXpert sputum (-) |
| 013 | TB-01-0137 | Not Detected | 0 | NA | Smear (-), Culture (-), GeneXpert sputum (-) |
| 014 | TB-01-0145 | Not Detected | 0 | NA | Smear (-), Culture (-), GeneXpert sputum (-) |

**Supplementary Table 5.** Fluorescence cutoff point derivation and associated sensitivities and specificities

| | Cutoff Value (FU) | Sensitivity (95% CI) | Specificity (95% CI) |
|---|---|---|---|
| **Mean + 3 SD of negative controls** | | | |
| Training set (n = 83) | 3287.03 | 0.93 (0.85, 1.00) | 1.00 (1.00, 1.00) |
| Test set (n = 32) | | 1.00 (1.00, 1.00) | 1.00 (1.00, 1.00) |
| **Youden index** | | | |
| Training set (n = 83) | 2895.42 | 0.96 (0.89, 1.00) | 1.00 (1.00, 1.00) |
| Test set (n = 32) | | 0.93 (0.80, 1.00) | 1.00 (1.00, 1.00) |
| **TPP 2024 sensitivity standard*** | | | |
| Training set (n = 83) | 7047.50 | 0.85 | 1.00 (1.00, 1.00) |
| Test set (n = 32) | | 0.82 (0.65, 1.00) | 1.00 (1.00, 1.00) |
| **TPP 2024 specificity standard** | | | |
| Training set (n = 83) | 2895.42 | 0.96 (0.89, 1.00) | 0.98 |
| Test set (n = 32) | | 0.93 (0.80, 1.00) | 1.00 (1.00, 1.00) |

*For a sputum-based near-POC test.

.

**Supplementary Table 6**. Primer, crRNA, and reporter sequences.

| Name | Label | Oligo type; company | Sequence | Gene assay name |
|---|---|---|---|---|
| IS6110_cr1 | crRNA | custom RNA; IDT | /AltR1/rUrArArUrUrUrCrUrArCrUrArArGrUrGrUrArGrArUrCrUrArCrCrCrArCrArGrCrCrGrGrUrUrArGrGrU/AltR2/ | Cas12_IS6110_cr1 |
| IS6110_cr2 | crRNA | custom RNA; IDT | /AltR1/rUrArArUrUrUrCrUrArCrUrArArGrUrGrUrArGrArUrArCrCrGrGrCrUrGrUrGrGrGrUrArGrCrArGrArC/AltR2/ | Cas12_IS6110_cr2 |
| IS6110_cr3 | crRNA | custom RNA; IDT | /AltR1/rUrArArUrUrUrCrUrArCrUrArArGrUrGrUrArGrArUrArArArGrArCrCrGrCrGrUrCrGrGrCrUrUrUrCrU/AltR2/ | Cas12_IS6110_cr3 |
| IS6110_cr4 | crRNA | custom RNA; IDT | /AltR1/rUrArArUrUrUrCrUrArCrUrArArGrUrGrUrArGrArUrCrCrGrCrGrGrGrUrGrGrUrCrCrCrGrGrArCrArG/AltR2/ | Cas12_IS6110_cr4 |
| IS6110_cr5 | crRNA | custom RNA; IDT | /AltR1/rUrArArUrUrUrCrUrArCrUrArArGrUrGrUrArGrArUrCrGrGrGrCrArCrCrGrUrArArArCrArCrCrGrUrA/AltR2/ | Cas12_IS6110_cr5 |
| IS6110_cr6 | crRNA | custom RNA; IDT | /AltR1/rUrArArUrUrUrCrUrArCrUrArArGrUrGrUrArGrArUrArArArUrCrGrCrGrUrUrCrGrCrCrCrUrUrCrGrC/AltR2/ | Cas12_IS6110_cr6 |
| IS1081_cr1 | crRNA | custom RNA; IDT | /AltR1/rUrArArUrUrUrCrUrArCrUrArArGrUrGrUrArGrArUrGrCrGrCrCrArGrArUrCrUrGrCrUrUrGrGrGrGrA/AltR2/ | Cas12_IS1081_cr1 |
| IS1081_cr3 | crRNA | custom RNA; IDT | /AltR1/rUrArArUrUrUrCrUrArCrUrArArGrUrGrUrArGrArUrGrCrCrArUrGrArUrCrGrArCrArCrUrUrGrCrGrA/AltR2/ | Cas12_IS1081_cr3 |
| IS1081_cr4 | crRNA | custom RNA; IDT | /AltR1/rUrArArUrUrUrCrUrArCrUrArArGrUrGrUrArGrArUrUrCrArCrArCrCrArArGrUrGrUrUrUrCrGrArCrC/ | Cas12_IS1081_cr4 |

| | | | | |
|---|---|---|---|---|
| | | | AltR2/ | |
| C13_IS6110_A | crRNA | custom RNA; IDT | /AltR1/rGrArUrUrUrArGrArCrUrArCrCrCrCrArArArArArCrGrArArGrGrGrGrArCrUrArArArArCrGrGrCrArUrCrGrArGrGrUrGrGrCrCrArGrArUrGrCrArCrCrGrUrCrG/AltR2/ | Cas13_IS6110_A |
| C13_IS6110_B | crRNA | custom RNA; IDT | /AltR1/rGrArUrUrUrArGrArCrUrArCrCrCrCrArArArArArCrGrArArGrGrGrGrArCrUrArArArArCrGrArCrGrArUrCrArArCrGrGrCrCrUrArUrArCrArArGrArCrCrGrArG/AltR2/ | Cas13_IS6110_B |
| C13_IS6110_sg5 | crRNA | custom RNA; IDT | /AltR1/rGrArUrUrUrArGrArCrUrArCrCrCrCrArArArArArCrGrArArGrGrGrGrArCrUrArArArArCrCrUrArCrGrGrUrGrUrUrUrArCrGrGrUrGrCrCrCrGrCrArArArGrUrG/AltR2/ | Cas13_IS6110_C |
| C13_IS1081_A | crRNA | custom RNA; IDT | /AltR1/rGrArUrUrUrArGrArCrUrArCrCrCrCrArArArArArCrGrArArGrGrGrGrArCrUrArArArArCrCrArArArGrCrUrUrUrCrCrArArGrUrCrGrCrArArGrUrGrUrCrGrArU/AltR2/ | Cas13_IS1081_A |
| C13_IS1081_B | crRNA | custom RNA; IDT | /AltR1/rGrArUrUrUrArGrArCrUrArCrCrCrCrArArArArArCrGrArArGrGrGrGrArCrUrArArArArCrGrCrArGrCrGrCrCrGrCrArArGrCrGrArGrCrUrGrArArCrGrCrGrCrA/AltR2/ | Cas13_IS1081_B |
| C13_IS1081_C | crRNA | custom RNA; IDT | /AltR1/rGrArUrUrUrArGrArCrUrArCrCrCrCrArArArArArCrGrArArGrGrGrGrArCrUrArArArArCrGrCrGrGrGrCrUrArCrCrGrCrGrArArCrGrCrArGrCrGrArUrGrArGrC/AltR2/ | Cas13_IS1081_C |
| LR5_crRNA_1 | crRNA | custom RNA; IDT | /AlTR1/rUrArArUrUrUrCrUrArCrUrArArGrUrGrUrArGrArUrArCrArArArGrCrArCrArUrCrUrUrGrCrArCrCrGrC/AlTR2/ | Cas12_internalControl_1 |
| LR5_crRNA_2 | crRNA | custom RNA; IDT | /AlTR1/rUrArArUrUrUrCrUrArCrUrArArGrUrGrUrArGrArUrArArGrGrArGrCrArUrGrCrUrGrCrCrUrUrCrArArG/AlTR2/ | Cas12_internalControl_2 |
| LR5_crRNA_3 | crRNA | custom RNA; IDT | /AltR1/rUrArArUrUrUrCrUrArCrUrArArGrUrGrUrArGrArUrArArArGrCr | Cas12_internalControl_3 |

| | | | | |
|---|---|---|---|---|
| | | | ArCrArGrCrArCrUrUrArArUrCrCrU/AltR2/ | |
| IS6110_cr1-2_F | Primer | custom DNA; IDT | CGTAGGCGAACCCTGCCCAGGTCGACACATAGG | Cas12_IS6110_cr1 & Cas12_IS6110_cr2 |
| IS6110_cr1-2_R | Primer | custom DNA; IDT | CGATCTCGTCCAGCGCCGCTTCGGACCACCA | Cas12_IS6110_cr1 & Cas12_IS6110_cr2 |
| IS6110_cr3_F | Primer | custom DNA; IDT | AATTAGCGTGCTGGCCGGTCGAGCTCGGCC | Cas12_IS6110_cr3 |
| IS6110_cr3_R | Primer | custom DNA; IDT | GGGACAACGCCGAATTGCGAAGGGCGAACG | Cas12_IS6110_cr3 |
| IS6110_cr4_F | Primer | custom DNA; IDT | TGTGGCCGGATCAGCGATCGTGGTCCTGCGGGCTT | Cas12_IS6110_cr4 |
| IS6110_cr4_R | Primer | custom DNA; IDT | AGATGCACCGTCGAACGGCTGATGACCAAACTCGG | Cas12_IS6110_cr4 |
| IS6110_cr5_F | Primer | custom DNA; IDT | TGGCCACCTCGATGCCCTCACGGTTCAGGGTTAGCCACAC | Cas12_IS6110_cr5 |
| IS6110_cr5_R | Primer | custom DNA; IDT | GCGAACTCAAGGAGCACATCAGCCGCGTCCACGCCGCCAA | Cas12_IS6110_cr5 & Cas13_IS6110_C |
| IS6110_cr6_F | Primer | custom DNA; IDT | AGCTCGGCCGCGAAGAAAGCCGACGCGGTC | Cas12_IS6110_cr6 |
| IS6110_cr6_R | Primer | custom DNA; IDT | TGAAGCGCTTGCGGCGGGACAACGCCGAAT | Cas12_IS6110_cr6 |
| IS1081_cr1_F | Primer | custom DNA; IDT | ACCTCTCGGTTGAGGCGTTCCTGGGGGTTGTTGGACCAGA | Cas12_IS1081_cr1 |
| IS1081_cr1_R | Primer | custom DNA; IDT | ACCTCGACACCGCCCGCACCGACCTGCTGGCGTTCACCGC | Cas12_IS1081_cr1 |
| IS1081_cr3_F | Primer | custom DNA; IDT | GGTCCGAAACGCCTCTACGGCTTCGTCGAGCTCTT | Cas12_IS1081_cr3 |
| IS1081_cr3_R | Primer | custom DNA; IDT | CCTGGTCGAAACACTTGGTGTGACAAAGCTTTCCA | Cas12_IS1081_cr3 |
| IS1081_cr4_F | Primer | custom DNA; IDT | TTTGGCCATGATCGACACTTGCGACTTGGAAAGC | Cas12_IS1081_cr4 |

| | | | | |
|---|---|---|---|---|
| IS1081_cr4_R | Primer | custom DNA; IDT | ACCTGCTGGGAGTATCCACTCGCCGGATGGAGCGC | Cas12_IS1081_cr4 |
| C13_FwT7_IS6_A | Primer | custom DNA; IDT | gaaatTAATACGACTCACTATAgggGTCCCGGACAGGCCGAGTTTGGTCATCAGCCGTT | Cas13_IS6110_A |
| C13_FwT7_IS6_B | Primer | custom DNA; IDT | gaaatTAATACGACTCACTATAgggTCGATGGACCGCCAGGGCTTGCCGGGTTTGATCA | Cas13_IS6110_B |
| C13_FwT7_IS6_sg5 | Primer | custom DNA; IDT | gaaatTAATACGACTCACTATAgggCCACCTCGATGCCCTCACGGTTCAGGGTTAGCCA | Cas13_IS6110_C |
| C13_FwT7_IS1_A | Primer | custom DNA; IDT | gaaatTAATACGACTCACTATAgggCGCCTCTACGGCTTCGTCGAGCTCTTTGGCCATG | Cas13_IS1081_A |
| C13_FwT7_IS1_B | Primer | custom DNA; IDT | gaaatTAATACGACTCACTATAgggCCAGCAGGTAGCAGGTCGCCACCACGCTGGTCAG | Cas13_IS1081_B |
| C13_FwT7_IS1_C | Primer | custom DNA; IDT | gaaatTAATACGACTCACTATAgggTCACGGTGGCGGTAGCCGTTGCGCTGATTGGACC | Cas13_IS1081_C |
| C13_Rv_IS6_A | Primer | custom DNA; IDT | TTACGGTGCCCGCAAAGTGTGGCTAACCCTGAACCGTGAG | Cas13_IS6110_A |
| C13_Rv_IS6_B | Primer | custom DNA; IDT | TCGGAGCGGTCGGAAGCTCCTATGACAATGCACTAGCCGA | Cas13_IS6110_B |
| C13_Rv_IS6_B | Primer | Custom DNA; IDT | GCGAACTCAAGGAGCACATCAGCCGCGTCCACGCCGCCAA | Cas13_IS6110_C |
| C13_Rv_IS1_A | Primer | custom DNA; IDT | ACTCGCCGGATGGAGCGCCTGGTCGAAACACTTGGTGTGA | Cas13_IS1081_A |
| C13_Rv_IS1_B | Primer | custom DNA; IDT | CCAAGCTGCGCCAGGGCAGCTATTTCCCGGACTGGCTGCT | Cas13_IS1081_B |
| C13_Rv_IS1_C | Primer | custom DNA; IDT | TTCATCGCCGCCTTGATGGGGGCTGAAGCCGACGCCCTGT | Cas13_IS1081_C |
| LR5_1_FW | Primer | custom DNA; IDT | gaaacatgtgctgtgtcaactcagggttaaatggattaag | Cas12_internal Control_1 |
| LR5_1_RV | Primer | custom DNA; IDT | tggtgatgactcttaaggagcatgctgcctttcaagcatct | Cas12_internal Control_1 |
| LR5_2_FW | Primer | custom DNA; IDT | tggattaagggcggtgcaagatgtgctttgttaaacagat | Cas12_internal Control_2 |
| LR5_2_RV | Primer | custom DNA; IDT | tttgtgtccctgggtacttgagattagggagtggtgatga | Cas12_internal Control_2 |
| LR5_3_FW | Primer | custom DNA; IDT | gggcagcaatactgctttgtaaagcattgagatgtttatg | Cas12_internal Control_3 |
| LR5_3_RV | Primer | custom DNA; IDT | caaacacgtgaacaaaggtctttgcatcatagacaaggta | Cas12_internal Control_3 |

| | | | | |
|---|---|---|---|---|
| FAM-5C-Q | Reporter | custom DNA; IDT | /56-FAM/CCCCC/3IABkFQ/ | Cas12 fluorescent assays |
| HEX-5C-Q | Reporter | custom DNA; IDT | /5HEX/CCCCC/3IABkFQ/ | Cas12-13 dual fluorescence assays |
| FAM-6U-Q | Reporter | custom RNA; IDT | /56-FAM/rUrUrUrUrUrU/3IABkFQ/ | Cas13 and Cas12-13 fluorescence assays |
| FAM-14U-Bio | Reporter | custom RNA; IDT | /56-FAM/rUrUrUrUrUrUrUrUrUrUrUrUrUrU/3Bio/ | Cas13 LFAs |
| FAM-5C-Bio | Reporter | custom DNA; IDT | /56-FAM/CCCCC/3Bio/ | Cas12 LFAs |
| FAM-5C-Dig | Reporter | custom DNA; IDT | /56-FAM/CCCCC/3Dig_N/ | Cas12 LFAs |
| Bio-14C-FAM | Reporter | custom DNA; IDT | /5Biosg/CCCCCCCCCCCCCC/36-FAM/ | Cas12 LFAs |
| FAM-14C-Dig | Reporter | custom DNA; IDT | /56-FAM/CCCCCCCCCCCCCC/3Dig_N/ | Cas12 LFAs |

**Supplementary Table 7.** Gene Block Fragments Ordered

| Name | | Oligo type; company | Sequence | Relevant Figures |
|---|---|---|---|---|
| IS6110_A_target | Synthetic Fragment | gBlocks gene fragments; IDT | GAACCGGATCGATGTGTACTGAGATCCCCTATCCGTATGGTGGATAACGTCTTTCAGGTCGAGTACGCCTTCTTGTTGGCGGGTCCAGATGGCTTGCTCGATCGCGTCGAGGACCATGGAGGTGGCCATCGTGGAAGCGACCCGCCAGCCCAGGATCCTGCGAGCGTAGGCGTCGGTGACAAAGGCCACGTAGGCGAACCCTGCCCAGGTCGACACATAGGTGAGGTCTGCTACCCACAGCCGGTTAGGTGCTGGTGGTCCGAAGCGGCGCTGGACGAGATCGGCGGGACGGGCTGTGGCCGGATCAGCGATCGTGGTCCTGCGGGCTTTGCCGCGGGTGGTCCCGGACAGGCCGAGTTTGGTCATCAGCCGTTCGACGGTGCATCTGGCCACCTCGATGCCCTCACGGTTCAGGGTTAGCCACACTTTGCGGGCACCGTAAACACCGTAGTTGGCGGCGTGGACGCGGCTGATGTGCTCCTTGAGTTCGCCATCGCGCAGCTCGCGGCGGCTGGGCTCCCGGTTGATGTGGTCGTAGTAGGTCGATGGGGCGATCGGCACACCCAGCTCGGTCAGCTGTGTGCAGATCGACTCGACACCCCACCGCAAACCATCGGGGCCCTCGCGGTGGCCCTGATGATCGGCGATGAACCGGGTAATTAGCGTGCTGGCCGGTCGAGCTCGGCCGCGAAGAAAGCCGACGCGGTCTTTAAAATCGCGTTCGCCCTTCGCAATTCGGCGTTGTCCCGCCGCAAGCGCTTCAGCTCAGCGGATTCTTCGGTCGTGGTC | 2 A |
| IS6110_B_target | Synthetic Fragment | gBlocks gene fragments; IDT | AACCAGTCGACCCAGCGCGCGGTGGCCAACTCGACATCCTCGATGGACCGCCAGGGCTTGCCGGGTTTGATCAGCTCGGTCTTGTATAGGCCGTTGATCGTCTCGGCTAGTGCATTGTCATAGGAGCTTCCGACCGCTCCGACCGACGGTT | 2 A |
| IS1081_C,B_target | Synthetic Fragment | gBlocks gene fragments; IDT | TCCGGCGAGTGGATACTCCCAGCAGGTAGCAGGTCGCCACCACGCTGGTCAGTGCGCGTTCAGCTCGCTTGCGGCGCTGCAGCAGCCAGTCCGGGAAATAGCTGCCCTGGCGCAGCTTGGGGATCGCGACGTCGATGGTTGCGGCACGGGTGTCGAAATCACGGTGGCGGTAGCCGTTGC | 1 B, C and D; 2 A and B |

| | | | | |
|---|---|---|---|---|
| | | | GCTGATTGGACCGCTCATCGCTGCGTTCGCGGTAGCCCGCCCCGCACAGGGCGTCGGCTTCAGCCCCCATCAAGGCGGCGATGAACGTCGAGAGCAGCCCGC | |
| LR5_target_3 | Synthetic Fragment | gBlocks gene fragments; IDT | ggtgggacatgcgggcagcaatactgctttgtaaagcattgagatgtttatgtgtatgcatatctaaaagcacagcacttaatcctttaccttgtctatgatgcaaagacctttgttcacgtgtttgtctgctga | 3 B; Supplemental 4 E |
| LR5_target_1-2 | Synthetic Fragment | gBlocks gene fragments; IDT | cccgtgctctctgaaacatgtgctgtgtcaactcagggttaaatggattaagggcggtgcaagatgtgctttgttaaacagatgcttgaaggcagcatgctccttaagagtcatcaccactccctaatctcaagtacccagggacacaaaaactgcggaagg | Supplemental 4 C and D |

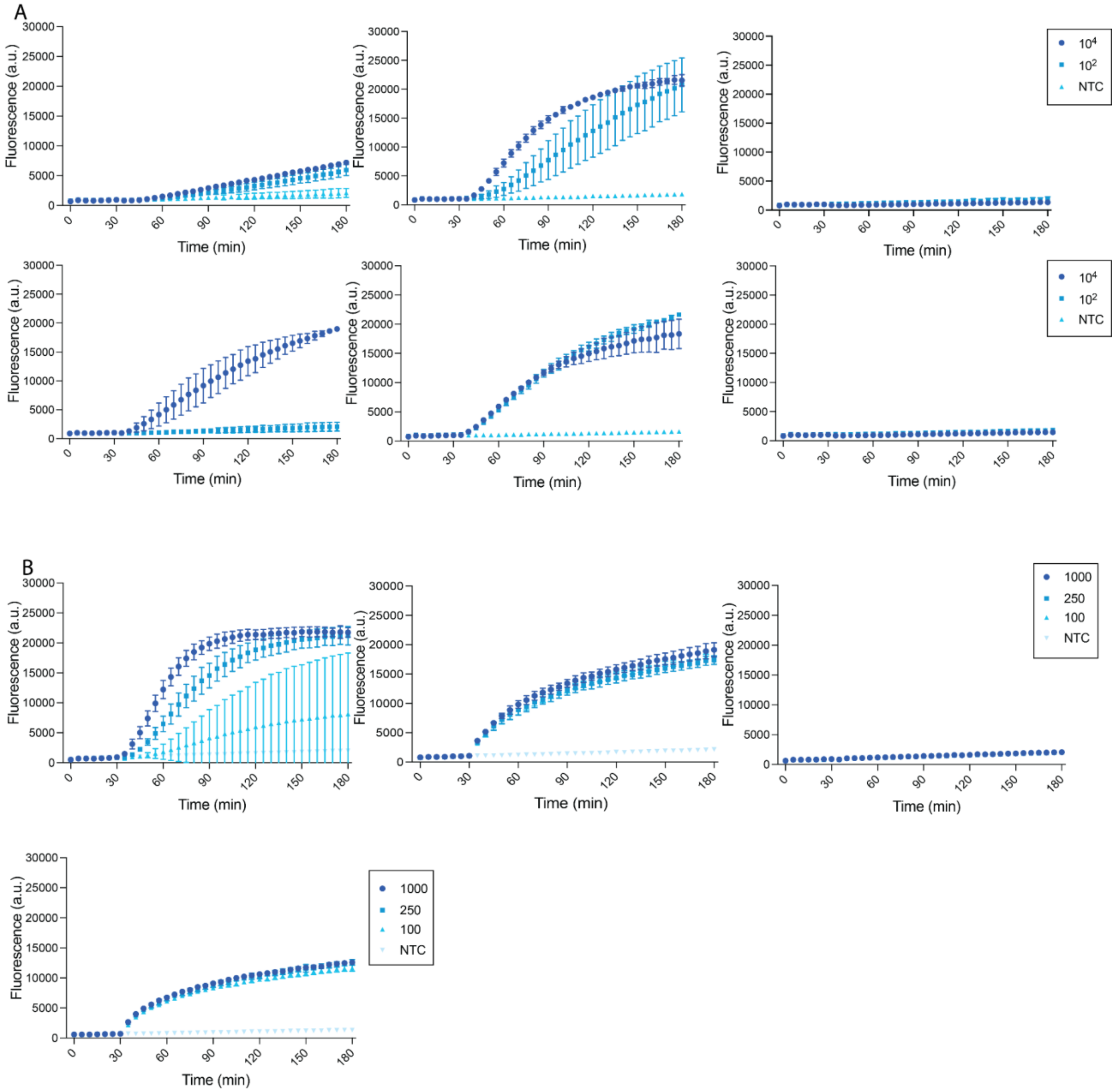


**Supplementary Figure 1.** Kinetics for panels seen in Fig. 1b and 1c. All units are in copies/µL. **A.** Kinetics from Fig. 1b. Target is synthetic DNA. cr1 (top left), cr2 (top middle), cr3 (top right), cr4 (bottom left), cr5 (bottom middle), cr6 (bottom right). **B.** Kinetics from Fig. 1c. Target is synthetic DNA and design cr5. 4 nM (top left), 4 nM spiked (top middle), 20 nM (top right), and 20 nM spiked (bottom left). In a, error bars: SD based on n=2 replicates. In b, error bars: SD based on n=3 technical replicates.

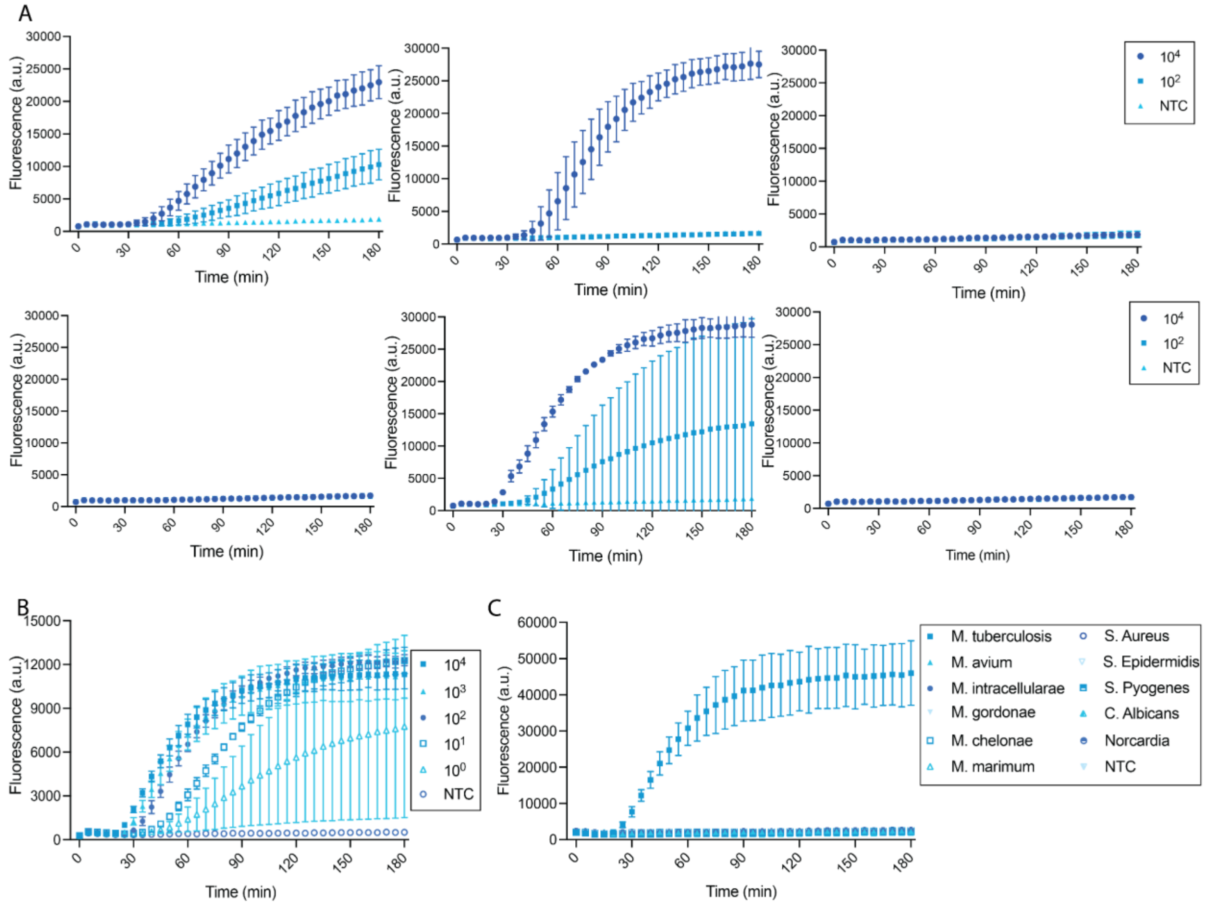


**Supplementary Figure 2.** Kinetics for panels seen in Fig. 1d, 1e and 1f. All units are in copies/µL. **A**. Kinetics from Fig. 1d. Target is synthetic DNA. cr1 (top left), cr2 (top middle), cr3 (top right), cr4 (bottom left), cr5 (bottom middle), cr6 (bottom right). **B.** Kinetics from Fig. 1e. Target is H37Rv. **C.** Kinetics from Fig. 1f. In a, error bars: SD based on n=2 technical replicates. In b and c, error bars: SD based on n=3 technical replicates.

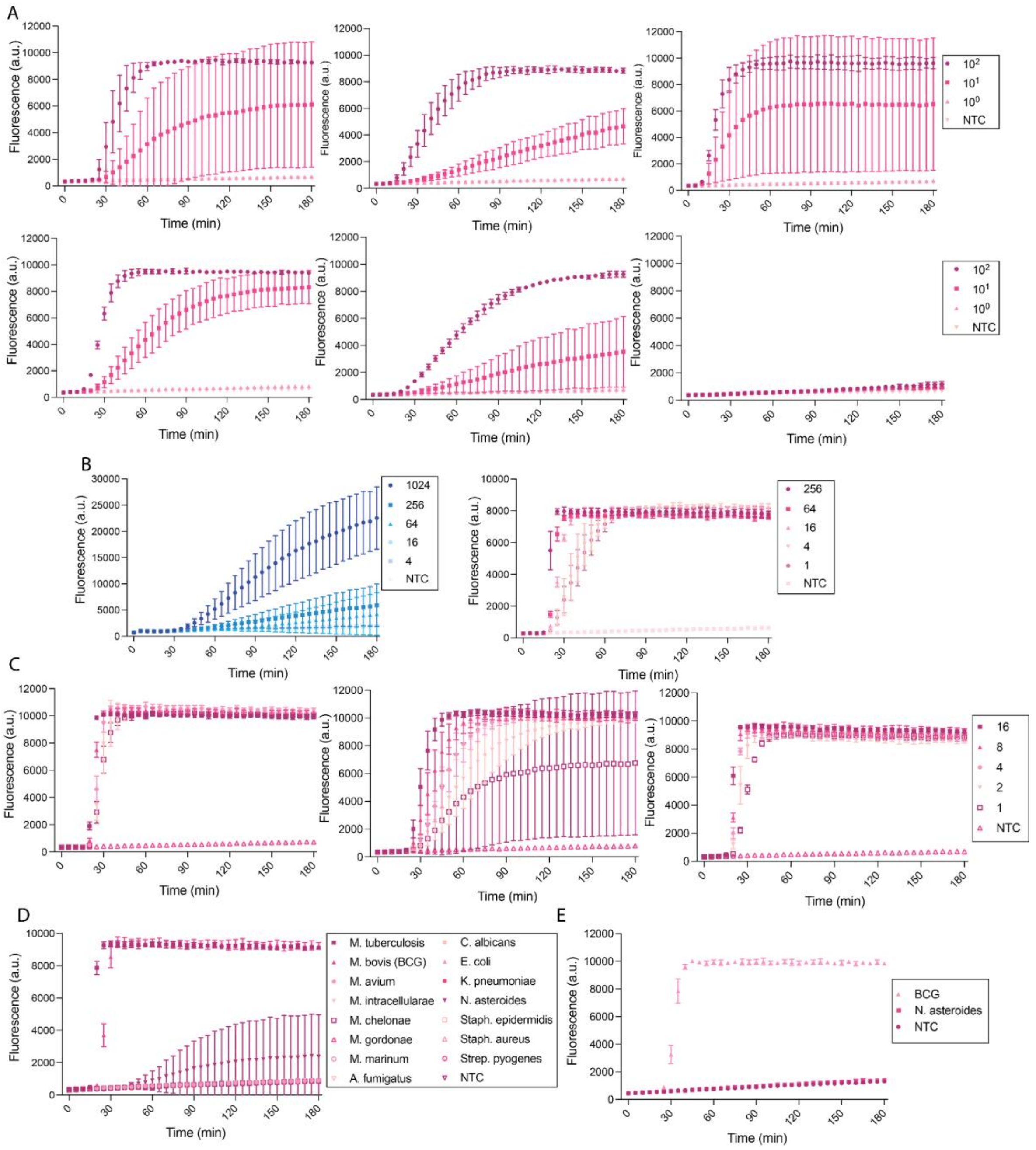


**Supplementary Figure 3.** Kinetics related to Fig. 2. All units are in copies/µL. **A.** Kinetics from Fig. 2a. Target is synthetic DNA. IS6110 A (top left), IS6110 B (top middle), IS6110 C (top right), IS1081 A (bottom left), IS1081 B (bottom middle), IS1081 C (bottom right). **B.** Kinetics from Fig. 2b. Target is synthetic DNA. Cas12a kinetics for cr5 (left) and Cas13a kinetics for IS6110 C (right). **C.** Kinetics from Fig. 2c. IS6110 alone. Target is H37Rv. **D.** Kinetics from Fig. 2d. **E.** Dual-detection Cas13 kinetics for *N. asteroides* at 2.5 ng/µL. In all panels, error bars: SD based on n=3 technical replicates.

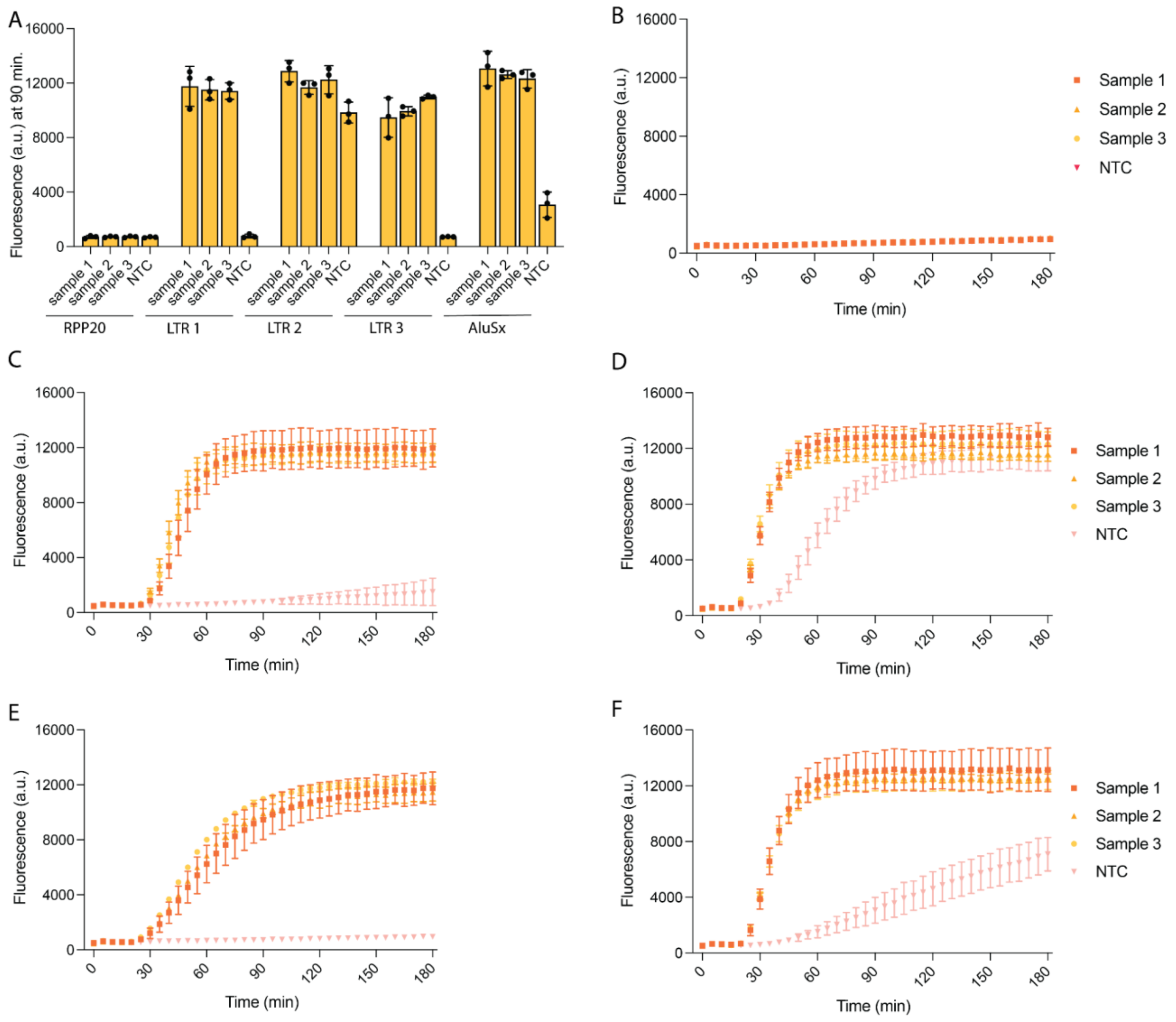


**Supplementary Figure 4.** Determining Internal Control Target. All samples used are saliva diluted 1:10 in nuclease free water. Cas12a is at a concentration of 20 nM. **A.** Guide designs targeting various human endogenous genes. RPP20 (ribonuclease P protein subunit p20), LTR 1-3 (various guides targeting Long Terminal Repeat of the endogenous retrovirus ERVK), and AluSx (alu element, transposable element). **B.** Kinetics for RPP20 (cross section shown in a). **C.** Kinetics for LTR (cross section shown in a). **D.** Kinetics for LTR 2 (cross section shown in a). **E.** Kinetics for LTR 3 (cross section shown in a). **F.** Kinetics for AluSx (cross section shown in a).
In all panels, error bars: SD based on n=3 technical replicates.

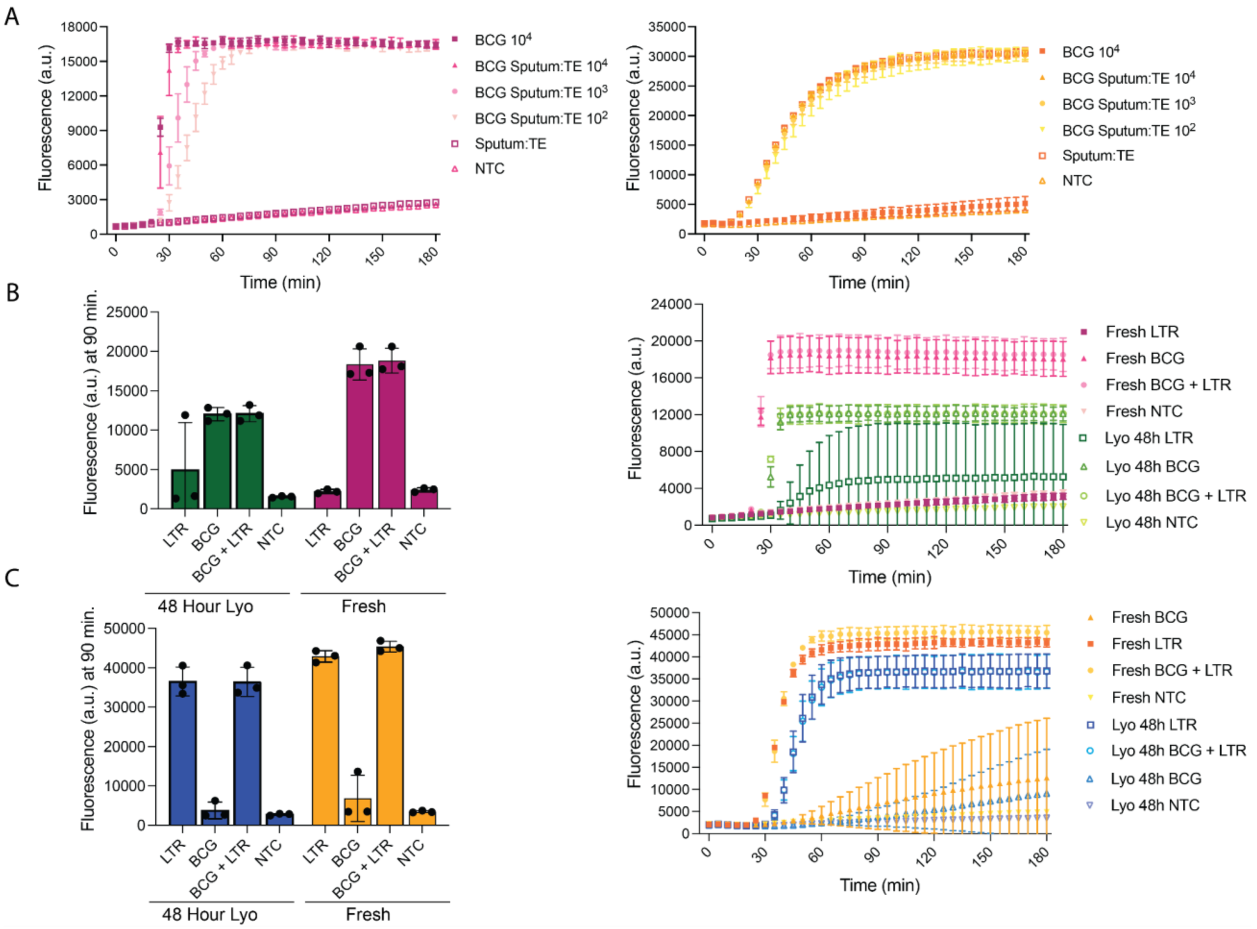


**Supplementary Figure 5.** Kinetics for panels seen in Fig. 3a and 3b. All units are in copies/µL. **A.** Kinetics from Fig. 3a. Cas13a *Mtb* (left) and Cas12a internal control (right). **B.** Supplementary data for duplex-assay seen in Fig. 3b (left). Kinetics for data seen in 3b (right). LTR is at $10^4$ copies/µL and BCG is at $10^3$ copies/µL. All targets are synthetic DNA. **C.** Supplementary internal control data seen in Fig. 3b (left). Kinetics for internal control data shown in 3b (right). LTR is at $10^4$ copies/µL and BCG is at $10^3$ copies/µL. All targets are synthetic DNA. In all panels, error bars: SD based on n=3 technical replicates.

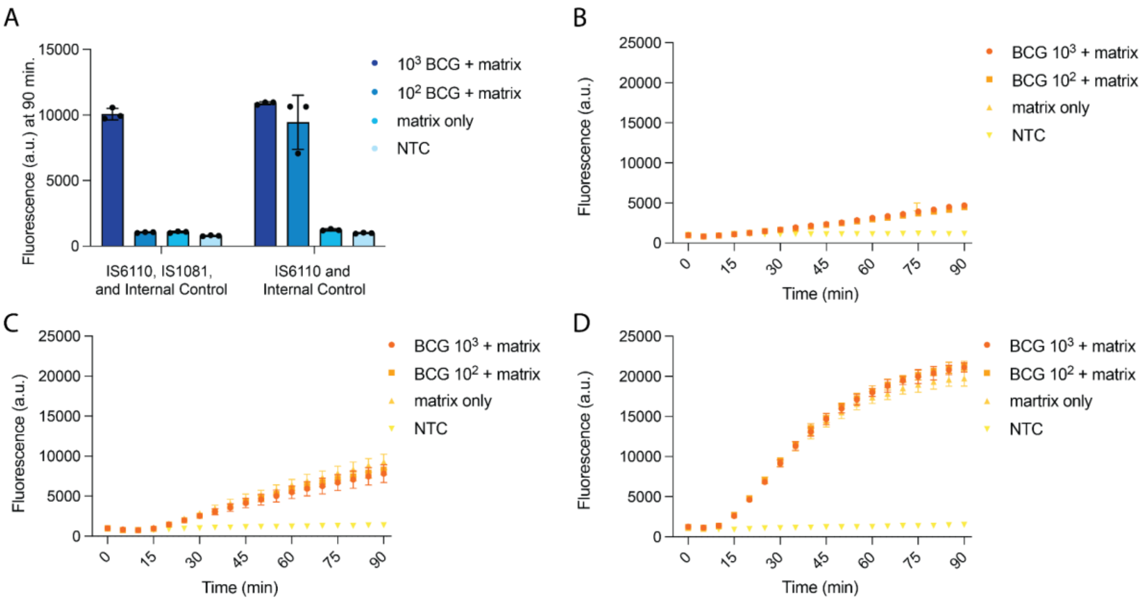


**Supplementary Figure 6**. Trialing assay configurations for single-, dual-, and triplex assays. **A.** Comparing performance of IS6110, IS1081, and internal control single-pot assay to IS6110 and Internal Control single-pot assay. Fluorescence detects the FAM channel and shows signals for BCG. Matrix is salivary sputum diluted 1:1. Fluorescence readout is 90 min. **B.** Cleavage kinetics of IS6110, IS1081, and internal control single-pot assay. Fluorescence detects the HEX channel and shows signals for the internal control. Matrix is salivary sputum diluted 1:1. **C.** Cleavage kinetics of IS6110 and internal control single-pot assay. Fluorescence detects the HEX channel and shows signals for the internal control. Matrix is salivary sputum diluted 1:1. **C.** Cleavage kinetics of internal control single-pot assay. Fluorescence detects the HEX channel and shows signals for the internal control. Matrix is salivary sputum 1:1. In all panels, error bars: SD based on n=3 technical replicates. In some cases, error bars are smaller than the data points.

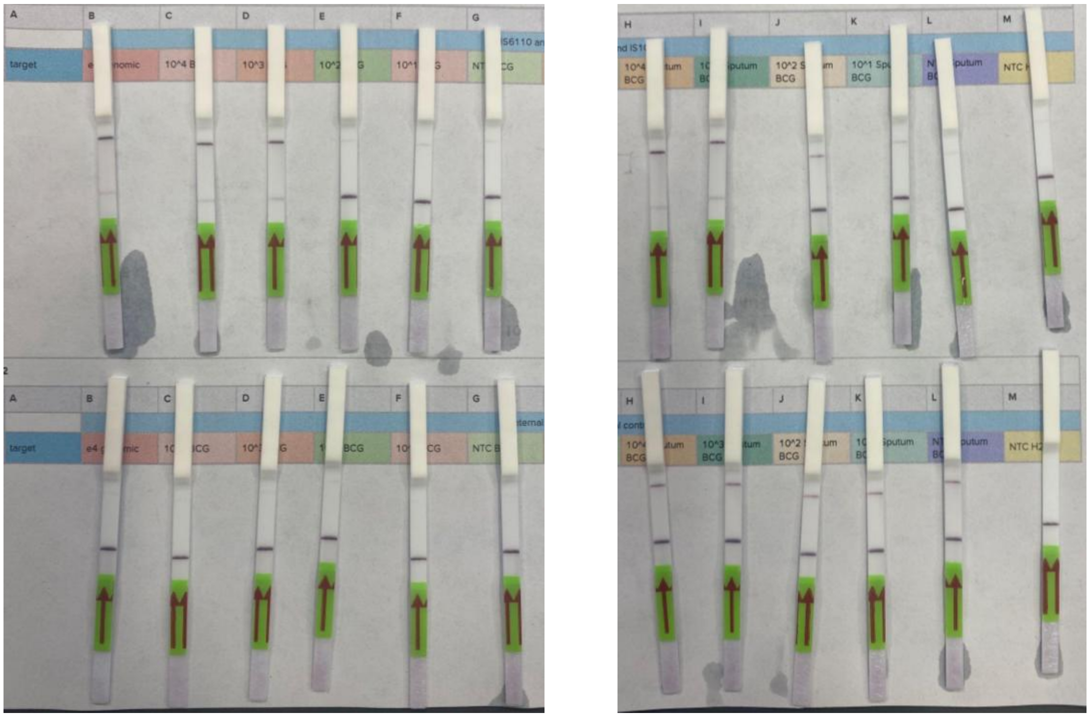

**Supplementary Figure 7.** Raw images of lateral flow strips seen in figure 3C. Cas13a detection (top) and Cas12a detection (bottom). BCG culture (left) and BCG culture in pooled sputum:TE (right).

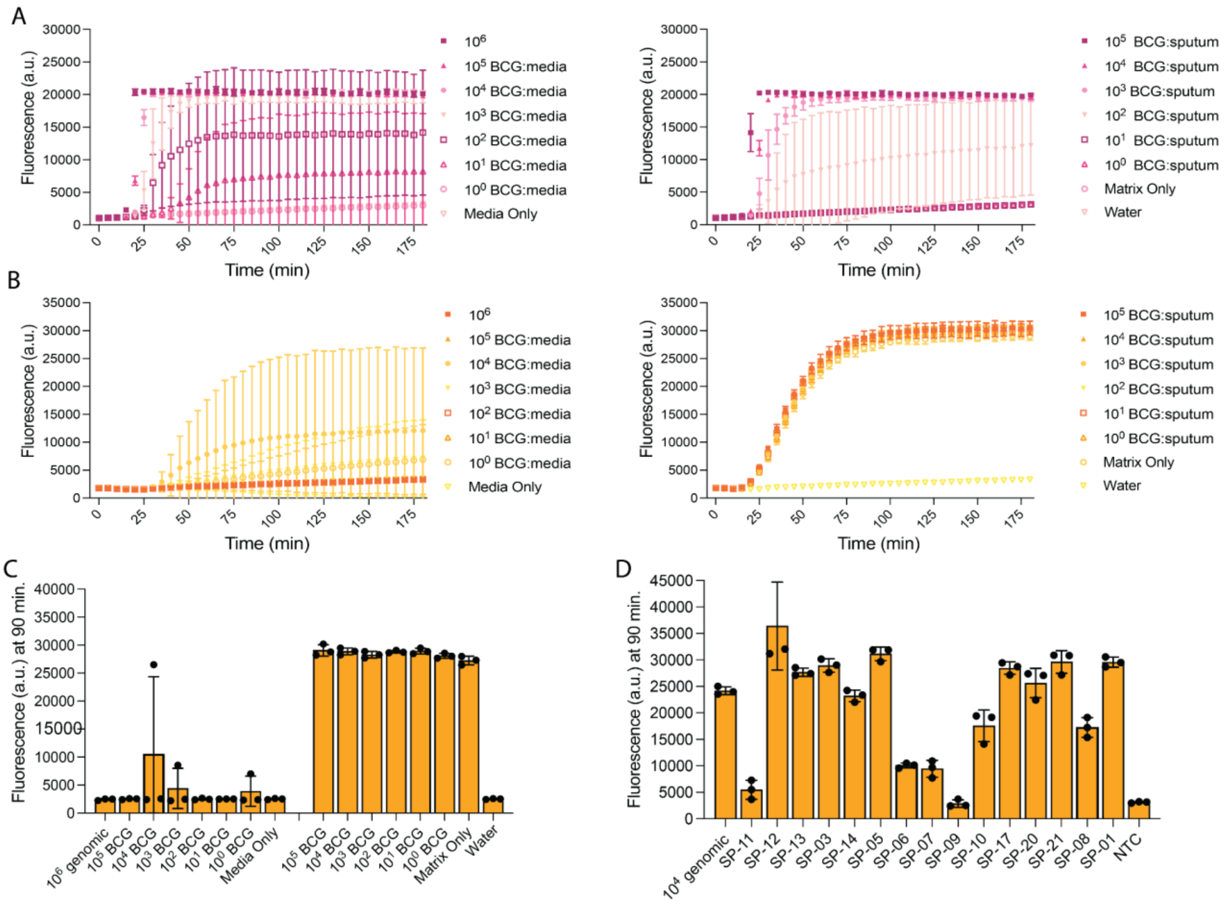

**Supplementary Fig. 8.** Internal Control and Kinetics Data for Fig. 3d and 3e. **A.** Cleavage connects for Cas13a in Fig. 3d. Fluorescence detects the FAM channel. **B.** Cleavage kinetics for internal control Cas12a for Fig. 3d. Fluorescence detects the HEX channel. **C.** Cas12a internal control data from Fig. 3d. Fluorescence detects the HEX channel. **D.** Cas12a internal control data from Fig. 3e. In all panels, error bars: SD based on n=3 technical replicates.

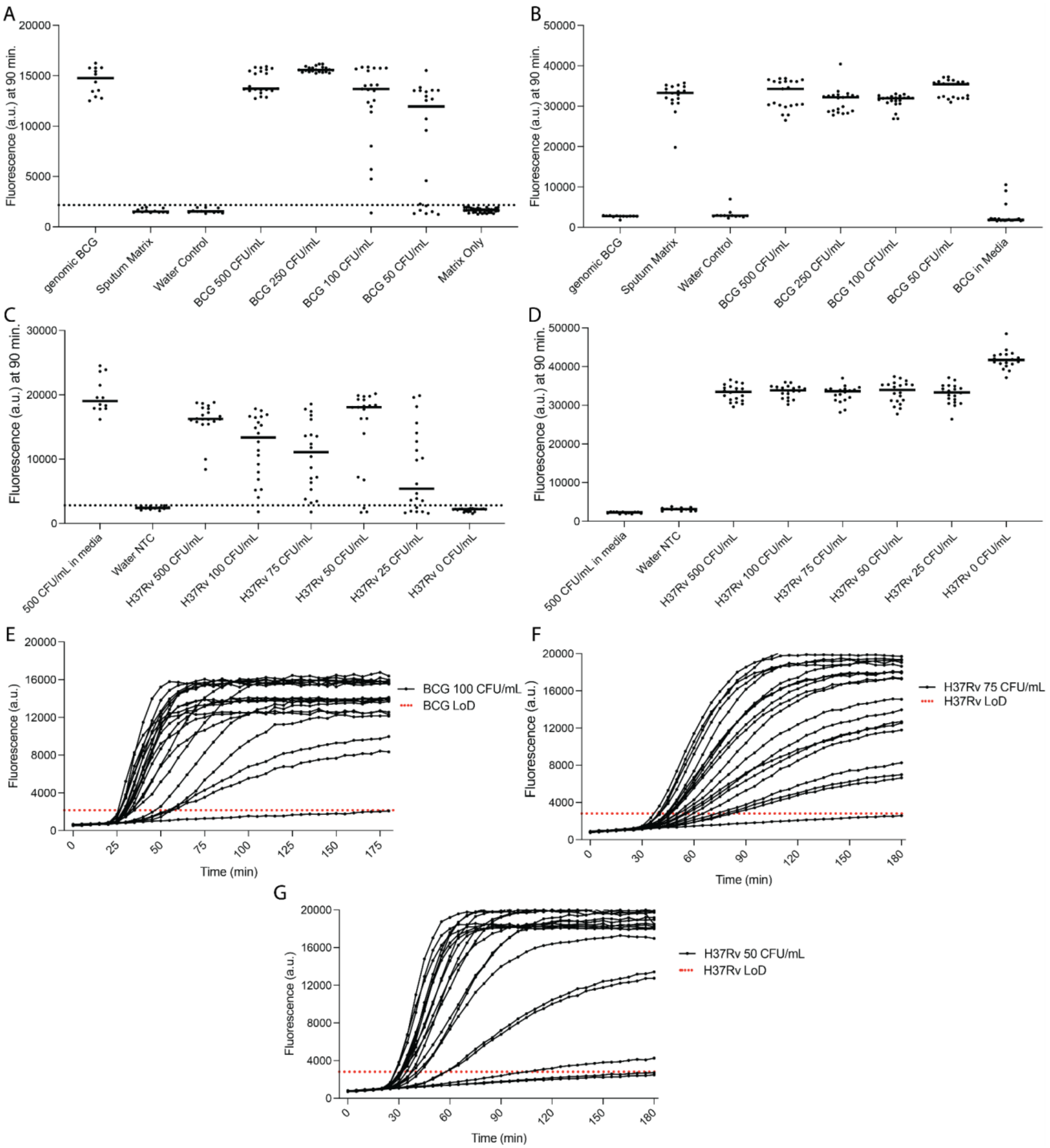


**Supplementary Figure 9.** Additional data for Fig. 3 for LoD. All units are in CFU/mL. All Cas13a assays are dual-detection. **A.** Individual fluorescence values from Fig. 3g, H37Rv LoD. Cas13a *Mtb*. **B.** Individual fluorescence values from Fig. 3g, H37Rv LoD. Cas12a internal control. **C.** Individual fluorescence values from Fig. 3f, H37Rv LoD. Cas13a *Mtb*. **D.** Individual fluorescence values from Fig. 3f, H37Rv LoD. Cas12a internal control. **E.** Kinetics from Fig. 3g, BCG LoD. Cas13a *Mtb* detecting 100 CFU/mL. **F.** Kinetics from Fig. 3f, H37Rv LoD. Cas13a *Mtb* detecting 75 CFU/mL. **G.** Kinetics for Cas13a detecting 50 CFU/mL of H37Rv.

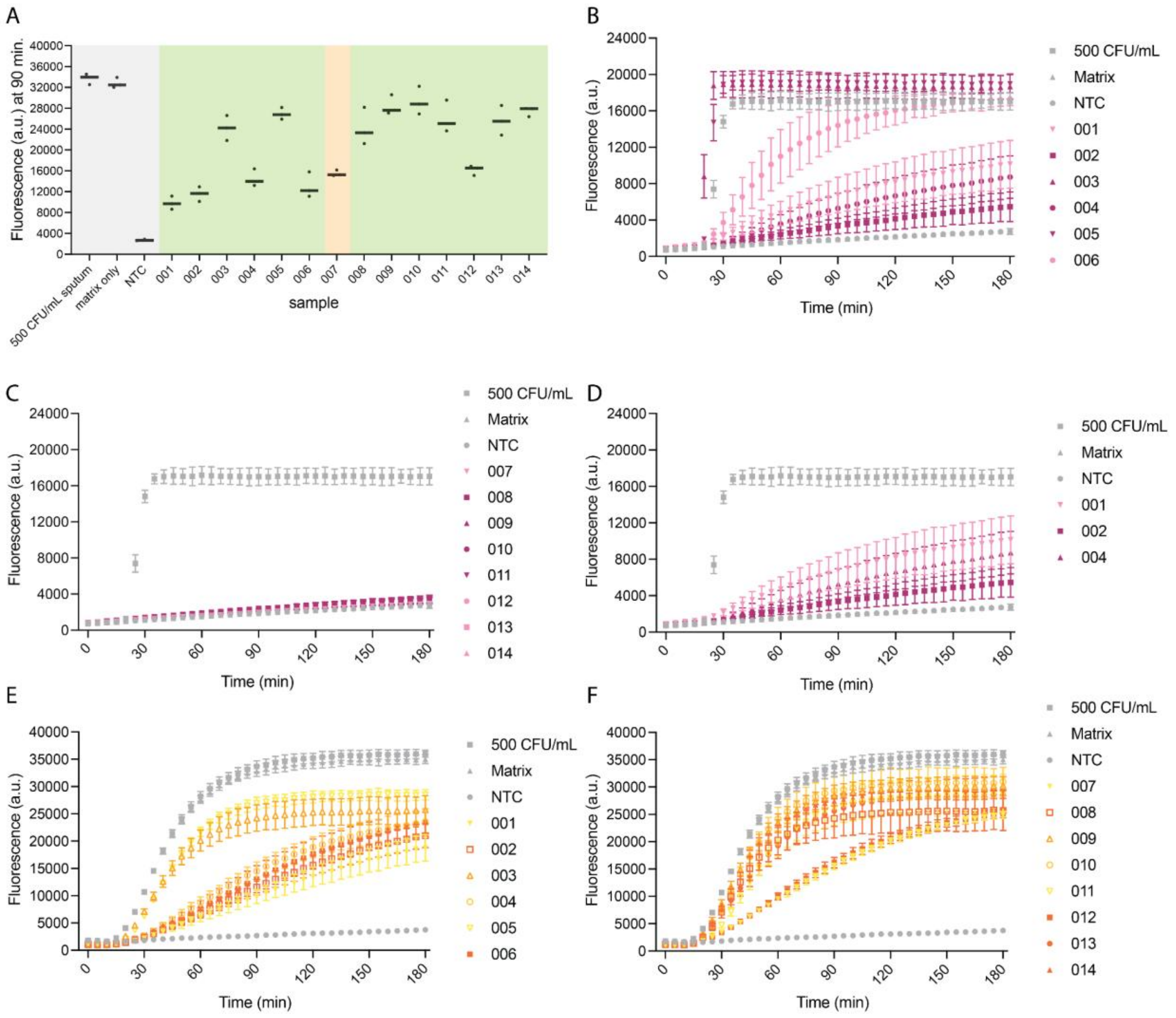


**Supplementary Figure 10.** Internal control and kinetics data seen in Fig. 4. **A.** Internal control data for Fig. 4b. **B.** Cas13a kinetics data for positive samples seen in Fig. 4b. **C.** Cas13a kinetics data for negative samples seen in Fig. 4b. **D.** Cas13a kinetics data for lowest signal positive samples seen in Fig. 4b. **E.** Internal control data for positive samples seen in Fig. 4b. **F.** Internal control data for negative samples seen in Fig. 4b. In b-f, error bars: SD based on n=3 technical replicates.
